# Intra-cellular accumulation of amyloid is a marker of selective neuronal vulnerability in Alzheimer’s disease

**DOI:** 10.1101/2023.11.23.23298911

**Authors:** Alessia Caramello, Nurun Fancy, Clotilde Tournerie, Maxine Eklund, Vicky Chau, Emily Adair, Marianna Papageorgopoulou, Johanna Jackson, John Hardy, Paul M. Matthews

## Abstract

Characterisation of vulnerable neurons that die earliest with Alzheimer’s disease (AD) could provide rationale treatment targets to slow or prevent neurodegeneration. We used imaging mass cytometry to identify the neuronal sub-types lost earliest in AD and explored associated mechanisms with paired single nuclear transcriptomics in *post-mortem* middle temporal gyri from diseased donors. We found L5-6 RORB^+^ and L3-6 GAD1^+^ neurons show the greatest loss in AD. These neuronal subtypes also accumulated β-amyloids intracellularly. By contrast, pTau^+^ tangles progressively formed in a distinct L3 RORB^+^GPC5^+^ subtype that appeared resilient. Both astrocytes and microglia expressed reactive phenotype markers with AD and reactive microglia were spatially associated with vulnerable neurons. RORB^+^ neuronal sub-types accumulating either amyloids or pTau showed increased expression of autophagy-related genes. In conclusion, we identified layer- and neuronal subtype-specific loss with AD that suggest intrinsic autophagy-related defects associated with intracellular accumulation of β-amyloids, rather than pTau, may initiate early neurodegeneration.

## INTRODUCTION

Selective degeneration and loss is observed in a subset of vulnerable neurons at early stages of Alzheimer’s disease (AD)^1^. Previous immunohistological studies suggested these include subtypes of excitatory (L2 reelin^+^ in the entorhinal cortex [EC]^2^ and L3-5 SMI32^+^ in the prefrontal cortex [PFC]^3^) and inhibitory neurons (SST^+^ and Calretinin^+^ in the piriform cortex^4^). PVALB^+^ and CALB1^+^ inhibitory neurons appear resilient to neurodegeneration^5^. Recently, single cell or nuclei transcriptomic studies suggested selective loss of GAD1^+^, LHX6^+^, SST^+^ and NPY^+^ inhibitory neurons in the PFC^6^, and RORB^+^ and CDH9^+^ excitatory neurons in the EC^7^. While this method may provide insights concerning neuronal molecular pathology associated with vulnerability, it is subject to multiple confounds for quantitation of cells and the primary identification of vulnerable neurons. Identifying the intrinsic characteristics underlying neuronal vulnerability and the mechanisms responsible for their selective loss could lead to rational designs of cell type-targeted diagnostics and novel approaches to treatment.

Neuronal vulnerability has often been associated with intraneuronal accumulation of hyper-phosphorylated Tau (pTau) which forms neurofibrillary tangles (NFT) and can lead to synaptic disfunction, toxicity and neuronal death^8–10^. However, vulnerability to NFT accumulation in the PFC appears to be distinct from vulnerability to death, which predominantly occurs in NFT-free neurons instead^6^. Other work has suggested that accumulation of pTau may not impair neuronal function, at least initially^11^ and may instead facilitate neurons escape apoptosis^12,13^. These studies raise the possibility that NFT accumulation is not a primary driver of neuronal death in AD but may be a marker of resilience.

An alternative hypothesis has been proposed that relates the accumulation of intraneuronal amyloid-β peptides, the primary constituent of extracellular amyloid plaques,^14^ with selective vulnerability. Aβ-42 is recognised to accumulate intra-neuronally in AD^15^. This accumulation is associated with impaired synaptic functions^16^ and neurotoxicity^17^. Intraneuronal Aβ-42 (intraAβ) was found to accumulate in reelin^+^ vulnerable neurons of the EC in both rats and humans^18^. Recent work suggests intraAβ accumulation can arise with impairments of neuronal autophagy–lysosomal pathway^19,20^. GWAS studies have implicated these pathways in susceptibility to AD^21^, suggesting that they play roles early, before neuronal loss^22,23^. Mouse models and human Down’s syndrome pathology suggest that intraAβ accumulation precedes plaque formation^24,25^. Decreases in relative numbers of neurons with intraAβ as the number of plaques increases supports the hypothesis that the accumulation is associated with mechanisms of early neuronal death^26^. Targeting intraAβ accumulation in neurons would therefore represent an effective therapy design to tackle the disease at early stages, aimed at enhancing neuronal reliance and reduce neuronal loss. However, the specific neuronal subtypes selectively accumulating intraAβ are not well described, nor is the relationship between intraAβ correlation and selective vulnerability in AD.

Microglial and astrocyte activation are observed in early AD^27,28^. Microglial activation contributes to the clearance of amyloid-β, but chronic activation can lead to pathological pruning of neuronal synapses and neurotoxicity^29,30^. Microglia express the majority of AD risk genes^31^, such as *TREM2*, a membrane receptor involved in amyloid-β phagocytosis. Variants of this gene (*R62H*/*R47H*) reduce amyloid-β binding and increase AD risk up to ∼3.9^32^. Reactive microglia reactivity also has been observed near intraAβ neurons^33^, suggesting these might be an early trigger of neuroinflammation in AD. Activated microglia and amyloid-β also can lead to neurotoxic astrocyte phenotypes^34^.

In this study, we sought to identify the intrinsic cell characteristics, local pTau/Aβ pathology and glial reactivity associated with selective neuronal vulnerability in AD and how these are aggravated by TREM2 AD risk alleles. To do this, we established a panel of 31 antibodies for imaging mass cytometry (IMC) to generate a single cell multiplexed protein expression spatial dataset of excitatory and inhibitory neuronal subpopulations, glial cells, and AD pathology. We then characterised neuronal sub-type abundance in 43 *post-mortem* MTG samples from non-disease control, early (Braak III-IV) and late (Braak V-VI) AD samples, including many from donors bearing the TREM2 variants (*R62H*/*R47H*)^35^. We identified selective loss of L2,3 and L5,6 RORB^+^ and L3,5 and L6 GAD1^+^ populations with AD and found that these neuronal sub-type also selectively accumulated intraAβ with early pathology. pTau increased in a district L3 RORB^+^GCP5^+^ neuronal subtype. Activated microglia with AD were associated spatially with vulnerable neurons. Both relative numbers of reactive glia and loss of the same neuronal subtypes were increased with AD in brains from donors carrying *TREM2* risk variants. Finally, paired transcriptomic analyses revealed dysregulation of genes involved in APP processing and autophagy-lysosomal pathway in RORB^+^ neurons accumulating AD pathological proteins. Together, these studies provide evidence for a functional association between intraAβ accumulation and selective neuronal loss in AD and suggest that this arises from intrinsic autophagy-related defects triggering early neurodegeneration.

## MATERIALS and METHODS

### Samples cohort

This study was carried out in accordance with the Regional Ethics Committee and Imperial College Use of Human Tissue guidelines. Cases were selected based first on neuropathological diagnosis (non-disease control [NDC] or AD) from UK brain banks (London Neurodegenerative Diseases Brain Bank [King’s College London], Newcastle Brain Tissue Resource, Queen’s Square Brain Bank [University College London], Manchester Brain Bank, Oxford Brain Bank and South West Dementia Brain Bank [University of Bristol]. We excluded cases with clinical or pathological evidence for small vessel disease, stroke, cerebral amyloid angiopathy, diabetes, Lewy body pathology (TDP-43), or other neurological diseases. Where the information was available, cases were selected with a *post-mortem* delay of less than 49 hours. The final cohort (Table 1; Fig. 1.D) was formed of 12 non-diseased controls (Braak 0-II) and 31 early (Braak III-IV; 4 samples) and late (Braak V-VI; 27 samples) AD human *post-mortem* FFPE middle temporal gyrus (MTG) samples. Of these, 4 controls and 7 late AD donors carried the AD high-risk *R62H* TREM2 variant, while 2 controls and 6 late AD donors carried the *R47H* variant. For antibody optimisation, one prefrontal cortex (PFC) control sample (age range: 81-85; sex: male; PM delay: 48 hours) was obtained as an FFPE block from Parkinson’s UK Brain Bank (Imperial College London). All of the brain banks used have generic ethics committee approval to function as research tissue banks and therefore did not require additional (UK) researcher-specific ethics approval.

**Figure 1.**
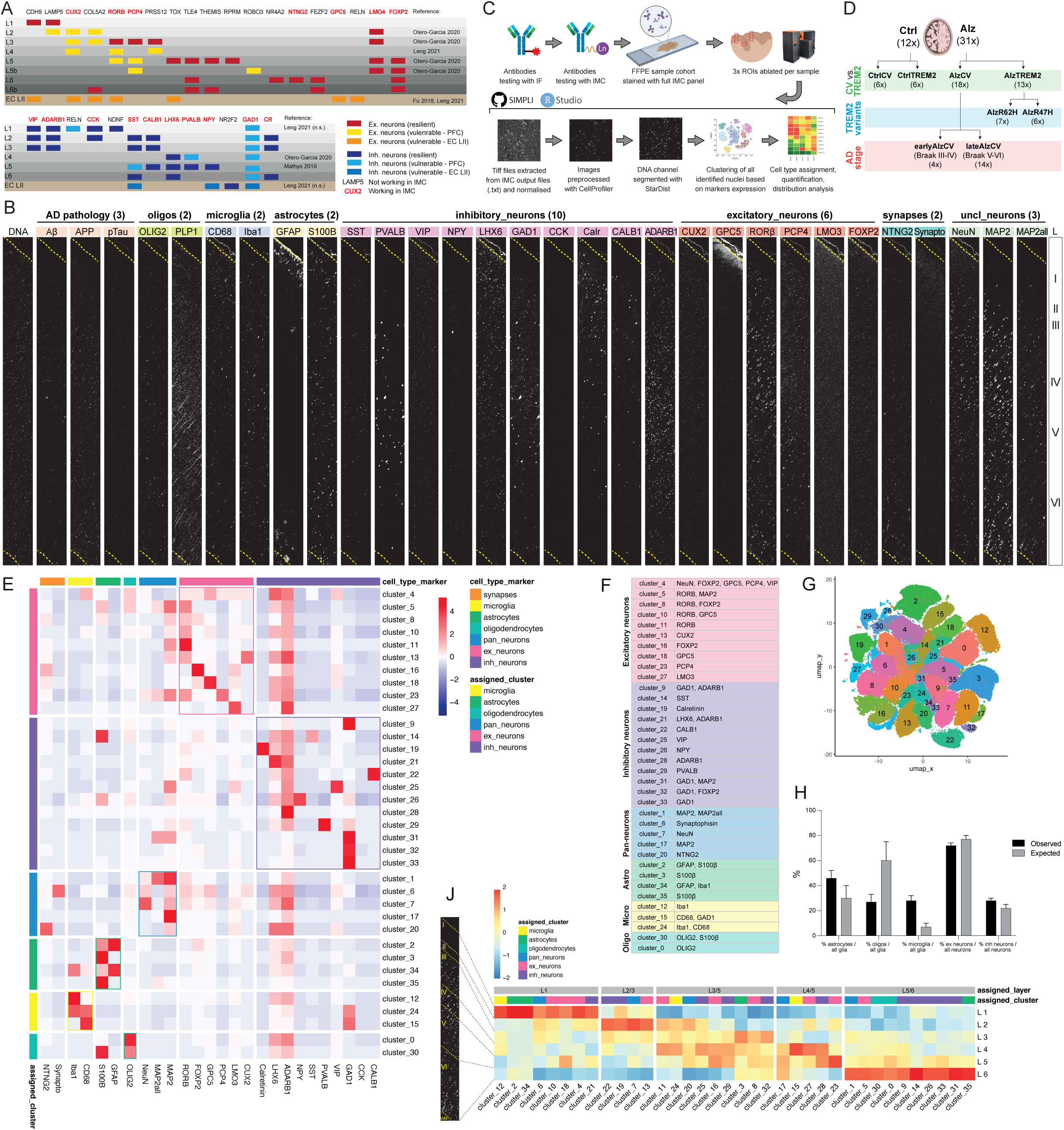
Establishment and analysis of IMC antibody panel for identification of neuronal and glial subtypes in human *post-mortem* brain (**A**) List of candidate neuronal markers tested for IMC antibody panel, with expected cortical distribution in MTG/PFC and EC LII, previous association with neuronal vulnerability and corresponding reference. Successful antibodies included in the final panel are indicated in red. (**B**) Example of IMC images obtained from one ROI processed with final antibody panel. Each ROI ablated span the entire length of the cortex (grey matter) from L1 to L6. (**C**) Schematic of whole methodological pipeline from antibodies testing (first IF then IMC), staining and ablation of full sample cohort, automated images analysis using SIMPLI and final data analysis in R. (**D**) Cohort of 12 non-disease controls and 31 AD cases processed. Depending on the analysis, these were divided based on expression of TREM common allele (CV) or risk variants (TREM2) and Braak stages. (**E**) Heatmap showing mean intensity of markers expression (columns) in each identified cluster (rows). Markers were grouped based on cell type specificity (coloured “cell_type_marker” bars on the top). Their intensity in each cluster was used to assign clusters to cell types (coloured “assigned_cluster” bars on the left). Coloured boxes highlight groups of cell type clusters expressing corresponding cell type markers. (**F**) Final assignment of neuronal and glial subtype to each clusters, with main markers expressed by each clusters indicated on the right. (**G**) UMAP plot of the 35 clusters identified from all detected cells. (**H**) Observed and expected proportion of astrocytes, oligodendrocytes and microglia to total glia population (left) and excitatory (which include pan-neuronal clusters) and inhibitory neurons to total neuronal population (right; CtrlCV samples only). (**J**) Distribution of cells from each cluster within the 6 cortical layers (CtrlCV samples only), normalised by total number of nuclei per layer. Cell type and layer assigned to clusters are indicated in the coloured and grey boxes above, respectively.

### Designing and testing of the neuronal antibody panel for IMC

An initial list of candidate markers for excitatory and inhibitory neuron and synapses, including vulnerable neurons, was identified screening previous literature that used single-nuclei RNAseq transcriptomic analyses, RNAscope and antibody-based stainings on fresh frozen or FFPE *post-mortem* human brain tissue^36–40^ to determine neuronal subtypes- and cortical layers-specific markers. The initial list was refined to prioritise markers shared among multiple publication and to have at least one marker per cortical layer or known neuronal subpopulation and include markers previously identified for vulnerable neurons^1,7,41^. To develop a panel applicable in different human brain regions, markers shared among EC, MTG and PFC were prioritised. Finally, antibodies commercially available in a carrier-free solution were also prioritised as a necessary condition for their conjugation to IMC metal isotopes. The initial list of 34 markers of neuronal subpopulations and synapses (Fig. 1.A; Table 2) was tested with immunofluorescence together with a known pan-neuronal marker (MAP2, MAP2all or NeuN) and with IMC upon conjugation to metal isotope. Of the initial list, 2 synapses (Synaptophysin, NTNG2) and 6 excitatory (CUX2, GPC5, RORB, PCP4, LMO3, FOXP2) and 10 inhibitory neurons markers (SST, PVALB, VIP, NPY, LHX6, GAD1, CCK, CR, CALB1, ADARB1) showed satisfactory signal from IMC to be used in the final panel (Fig. 1.A – markers in red).

To study glial cell activation and pathology development in the context of AD, the neuronal antibody panel was expanded with other antibodies already available in house: 3 pan-neuronal markers (NeuN, MAP2 and MAP2all, the latter covering also short isoforms of MAP2), 4 homeostatic and reactive astrocytic and microglial markers (S100β and GFAP, Iba1 and CD68, respectively), 2 oligodendrocytes and myelin markers (OLIG2 and PLP1), 3 AD pathology markers (β-amyloid, pTau and APP) and 1 nuclear marker (Intercalator). The final IMC panel designed (Table 3) included 31 antibodies, of which 2/3 are neuron-specific (Fig. 1.B).

### Immunostaining and confocal acquisition

The PFC control sample FFPE block used for antibody optimisation was sectioned at 8μm at the microtome and placed over super frost glass slides. Both locally sectioned and Brain Bank sections (also 8μm thick) were baked overnight at 60C for allowing FFPE sections to stick to the slide. Sections were then deparaffined and rehydrated with consecutive 5 minutes incubations in Histo-Clear II (2x), 100% ethanol (2x), 90% ethanol and 70% ethanol and finally washed in water for 5 minutes. Antigen retrieval was performed by incubating slides in EDTA pH9.0 for 20 minutes in a steam chamber, then cooled down for 10 minutes in ice and washed once in water and once in PBS for 5 minutes each. After applying a hydrophobic barrier around the tissue, slides were incubated in blocking buffer (10% donkey serum in PBS 0.3% Triton X-100) for an hour at room temperature inside a humid chamber. Primary antibodies were diluted (as indicated in Table 2) in blocking buffer and applied on slides overnight (for 1 or 2 nights) at 4C. Slides where then washed 3x in PBS before applying secondary antibodies and DAPI diluted 1:500 in blocking buffer and incubated for 1 hour at room temperature in a dark humid chamber. Finally, slides were washed twice with water for 5 minutes each, incubated with Sudan Black (1% in 70% ethanol) for 10 minutes in a dark humid chamber, washed with running tap water for 10 minutes and air dried. Cover slips were mounted with antifade. Stained sections were acquired at the Leica SP8 confocal microscope at 10x, 20x and 40x and images processed with ImageJ.

### IMC antibody conjugation, staining and acquisition

Primary antibodies that were not suspended in a carrier-free solution were first purified using antibody purification kits (mouse antibody kit, Protein A or G kit) based on the antibody type and specie and following manufacturer instructions (abcam; Table 2). Carrier-free and purified antibodies were then conjugated to lanthanide metals following Maxpar X8 protocol (Fluidigm, Standard BioTools, CA, USA).

Slides with FFPE sections were stained with the full conjugated antibody panel (Table 3) following the same protocol as for immunofluorescence. After the first night incubation with metal-tagged antibodies and iridium-intercalator, slides were washed with water 3x 10 minutes each and air dried for 20 minutes. IMC was performed using a Hyperion Tissue Imager (Fluidigm, Standard BioTools, CA, USA) coupled to a Helios mass cytometer. The instrument was first tuned using the manufacturer’s 3-Element Full Coverage Tuning Slide before the slides were loaded into the device. 3x ROIs were ablated per sample at a frequency of 200Hz with 1μm resolution. Each ROI was spanning the full cortical length from L1 to L6 (grey matter – representative ROI in Fig. 1.B), adding to a total of 6 hours of acquisition per sample (∼2h per ROI) keeping the total area acquired per sample equal among samples. Data was stored as .txt files that were used for subsequent processing (Fig. 1.C).

### Automated IMC image analysis

The SIMPLI^42^ pipeline was used for automated image processing and analysis (Fig. 1.C), which includes the following steps. First, SIMPLI performs image processing which includes extraction of single .txt files from each ROI to TIFF images and their normalisation and pre-processing using CellProfiler. Threshold smoothing scale, correction factor, lower and upper bounds, and manual threshold in the CellProfiler pipeline for image pre-processing was adjusted for each channel to remove background and keep specific signal only, as well as yield a total number of positive cells per channel that was comparable to manual counting (Fig. S1.A). All images were processed with the same CellProfiler pipeline. SIMPLI pixel-based analysis was then performed at this stage to quantify total Aβ^+^ and pTau^+^ pixel area (Fig. S4.C,G). Single-nuclei segmentation was then performed within SIMPLI based on the intercalator (191Ir/193Ir) channel using StarDist, with the “2D_versatile_fluo” model and a probability threshold of 0.05. Single-nuclei channels intensity was used by SIMPLI for masking all identified nuclei and identify all cells that expressed at least one of the markers used (the “all_cells” subset), apart from β-amyloids, pTau, APP and PLP1. To identify neuronal and non-neuronal subtypes, the “all_cells” subset underwent unsupervised clustering with Seurat (resolution 0.9) using again all neuronal and non-neuronal markers, but excluding β-amyloids, pTau, APP and PLP1 signals (Fig. 1.G).

### Assignment of cell types to clusters

Markers expressed within each cluster (Fig. 1.E) were used to assign clusters to neuronal/glial sub-types (Fig. 1.F): 10 clusters were assigned to excitatory neurons, 12 to inhibitory neurons, 5 represented pan-neurons (including synapses-only clusters), 4 astrocytes, 3 microglia and 2 oligodendroglial cells.

The largest cluster expressed OLIG2^+^ (cluster_0), which, together with a smaller OLIG2^+^S100B^+^ cluster (cluster_30), reflects the high abundance of oligodendroglial cells in the cortex. Immediately smaller in size, are the two main clusters for reactive GFAP^+^S100B^+^ (cluster_2) and non-reactive GFAP^-^S100B^+^ (cluster_30) astrocytes, also represented by two smaller similar clusters (cluster_34, cluster_35). Less abundant clusters were instead assigned to Iba1^+^ microglia representing their CD68^+^ activated (cluster_24) and CD68^-^ non-activated (cluster_12) status, including one cluster with co-localisation of CD68 and GAD1 signal (cluster_15).

Single neuronal subpopulations of excitatory and inhibitory neurons were identified for almost each of the neuronal markers used, with only RORB (cluster_5, cluster_8, cluster_10, cluster_11) and GAD1 (cluster_9, cluster_31, cluster_32, cluster_33) populations further sub-clustering into subpopulations co-expressing a different combination of markers. Pan-neuronal markers were weakly expressed in the neuronal sub-populations clusters, particularly NeuN. Conversely, clusters exclusively expressing pan-neuronal markers (including of the biggest clusters (cluster_1), together with cluster_7 and cluster_17) may identify neuronal subpopulations not represented by the combination of neuronal markers used. Apart from low ADARB1 and LHX6 expression, neuronal markers were mostly not expressed in glial clusters, indicating glial and neuronal markers mostly do not overlap and can detect these populations independently. Only clusters with strongest expression of ADARB1 and LHX6, and without glial markers, were considered as inhibitory and excitatory neurons. Similarly, markers for oligodendrocytes, astrocytes and microglia only showed little co-localisation with neuronal clusters, apart from S100B in cluster_14 (SST+). Cluster_4 was the only cluster expressing all excitatory neurons markers (RORB, FOXP2, GPC5, PCP4, LMO3, CUX2) and some inhibitory ones (ADARB1, LHX6, VIP). However, its location in L1 (Fig. S2) suggests it might represent unspecific signal from the meninges and was therefore excluded from all analyses.

### Data and statistical analyses

SIMPLI output data was further analysed using RStudio. Cell distribution within cortical layers was performed by analysing the Y coordinates of each identified nuclei: as all ROIs spanned the total length of the cortex (from beginning of L1 to end of L6), the Y coordinates represent the vertical location of nuclei in the cortex. A reference layers thickness^43^ was proportionally applied to all ROIs based on their maximum Y coordinate (representing total cortical thickness; Fig. 1.J). Aβ and pTau signal intensity was used to determine intra-cellular positivity for such markers in either the “all_cell” subset (Fig. S4.D,H), or in each identified cluster (Fig. 3.A,B), based on thresholds defined previously (Fig. S1.A). Plaque area and the 50μm surrounding ring were separately segmented using an ImageJ macro created in house. Cells location within the plaque or ring area were computed based on their XY coordinates (Fig. S5.J,K).

Statistical analysis was also performed in RStudio. Dirichlet analysis was applied for identifying changes in clusters proportions between healthy and diseased groups. Normal distribution of data was first tested using Shapiro-Wilk test. Analysis of groups variance and pair-group comparisons was then performed with ANOVA and Tukey tests or Kruskal–Wallis and Wilcoxon signed-rank test for normally and not normally distributed data, respectively. Only *p-adj* values were used for defining statistical difference between groups. Data expressed as percentage was transformed with *arcsin* before proceeding with normality and statistical tests. *p*-values are indicated as: non-significant, ns, p>0.05; *p≤0.05; **p≤0.01; ***p≤0.001; ****p≤0.0001.

### Single nucleus RNA transcriptomic dataset

The snRNAseq dataset used here was previously described^35^ and includes all samples used for the IMC experiment, plus 10 additional samples (3 controls and 1 AD sample carrying the common TREM2 variant [CtrlCV and AlzCV, respectively], control and 5 AD samples carrying TREM2 risk variants [CtrlTREM2 and AlzTREM2, respectively]; Fig. 5.A; Table 1). For this analysis, only the MTG-derived nuclei were used.

**Figure 2.**
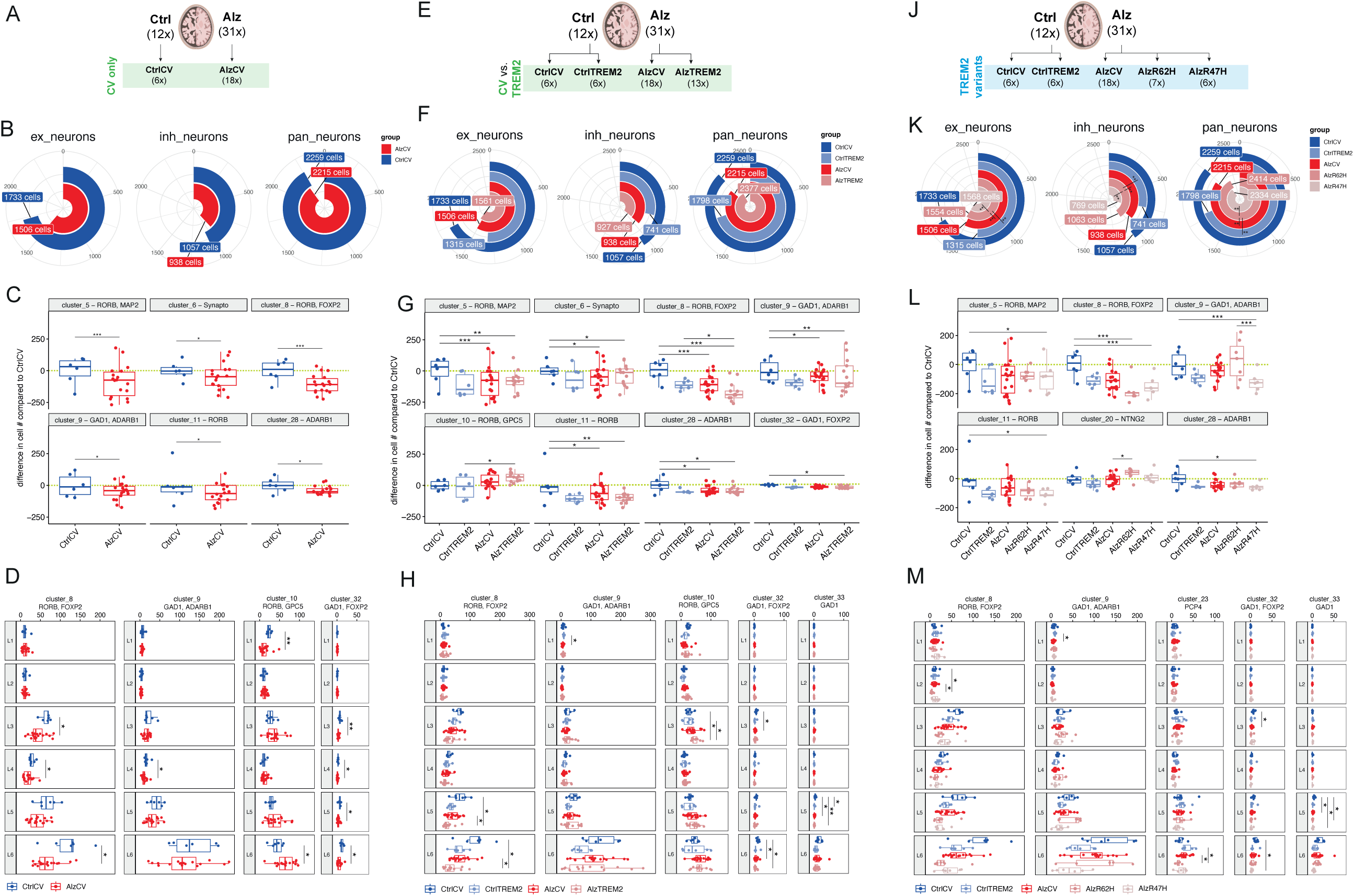
Neuronal subpopulations and cortical layers selectively affected in AD. (**A,E,J**) Schematic of the three samples grouping based on expression of TREM2 common allele or risk variant used for analyses shown in (B-D), (F-H) and (K-M), respectively. (**B,F,K**) Pie charts showing changes in total excitatory, inhibitory and pan-neuronal populations within sample groups. (**C,G,L**) Difference in average number of cells per cluster compared to CtrlCV. Yellow dashed line indicate the Y value of 0. (**D,H,M**) Distribution of nuclei along cortical layers (L1-6) from all clusters showing layer-specific vulnerabilities (distribution of all clusters shown in Fig. S2). Statistical significance was calculated with Dirichlet regression (C,G,L) or with ANOVA and Tukey tests or Kruskal–Wallis and Wilcoxon signed-rank test for normally and not normally distributed data respectively (D,H,M).

**Figure 3.**
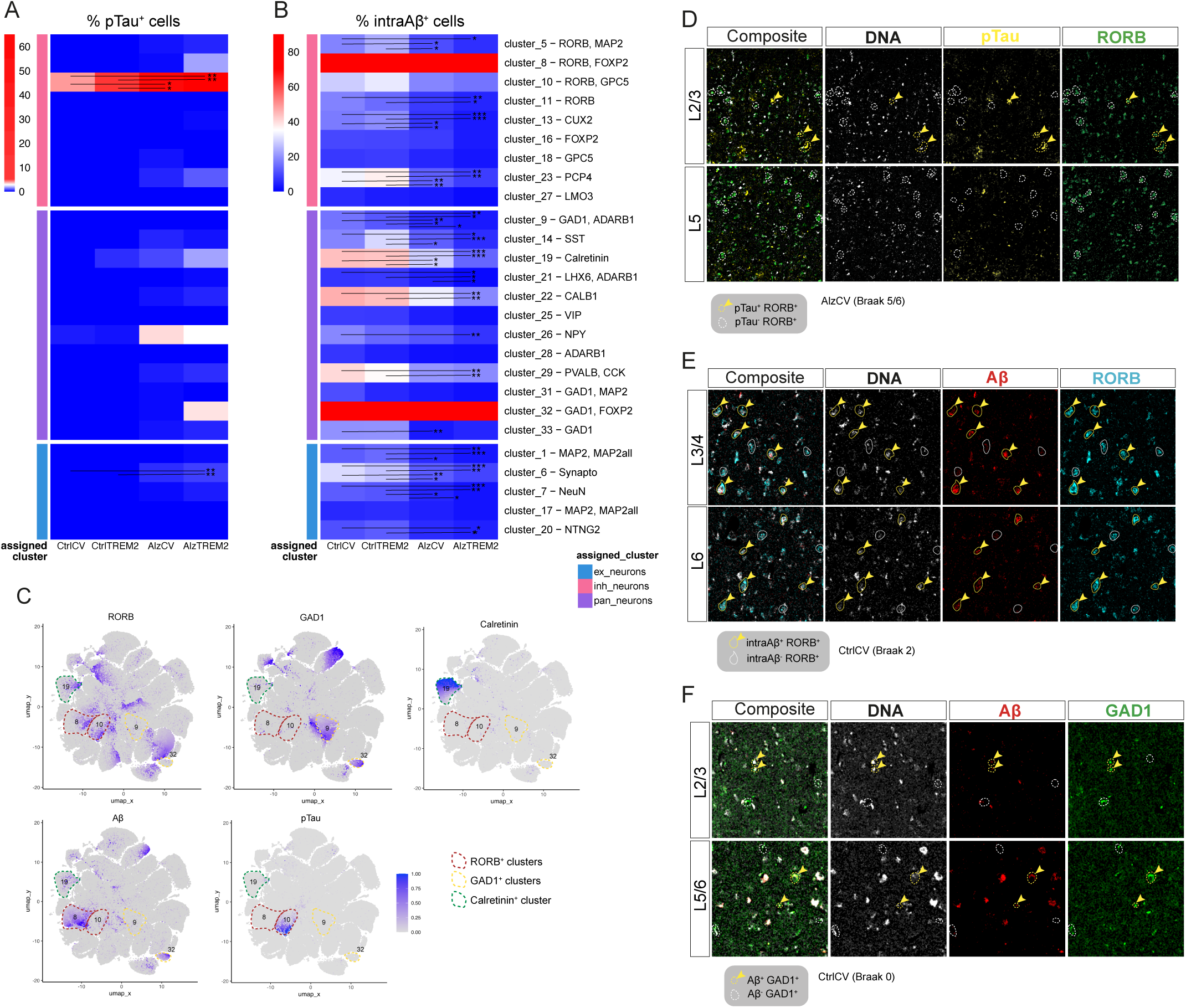
Intra-cellular pTau (NFT) and Aβ (intraAβ) accumulation in neuronal subtypes and its correlation to disease progression. (**A,B**) Proportion of pTau^+^ (A) and intraAβ^+^ (B) cells in each neuronal subtype cluster. Coloured boxes on the left highlight groups of neuronal types (excitatory, inhibitory and pan-neurons) to which clusters were assigned to (“assigned_cluster”). (**C**) UMAP plots of all clustered nuclei, highlighting expression of the four main neuronal markers (RORB, GAD1, Synaptophysin, Calretinin) accumulating AD pathological proteins (Aβ, pTau) as shown in (A,B). (**D**,**E**,**F**) IMC images showing co-localisation of: GAD1, Aβ in a CtrlCV (Braak 0) sample (E); RORB, Aβ in a CtrlCV (Braak 2) sample (F); RORB, pTau in a AlzCV (Braak 5/6) sample (G). Double-positive cells (yellow arrowheads) are found in the same layers as indicated from analyses in (Fig. 2.C,G,L). Statistical significance was calculated with ANOVA and Tukey tests or Kruskal–Wallis and Wilcoxon signed-rank test for normally and not normally distributed data respectively. Data expressed as percentage (A,B) was transformed with *arcsine* before proceeding with normality and statistical tests.

**Figure 4.**
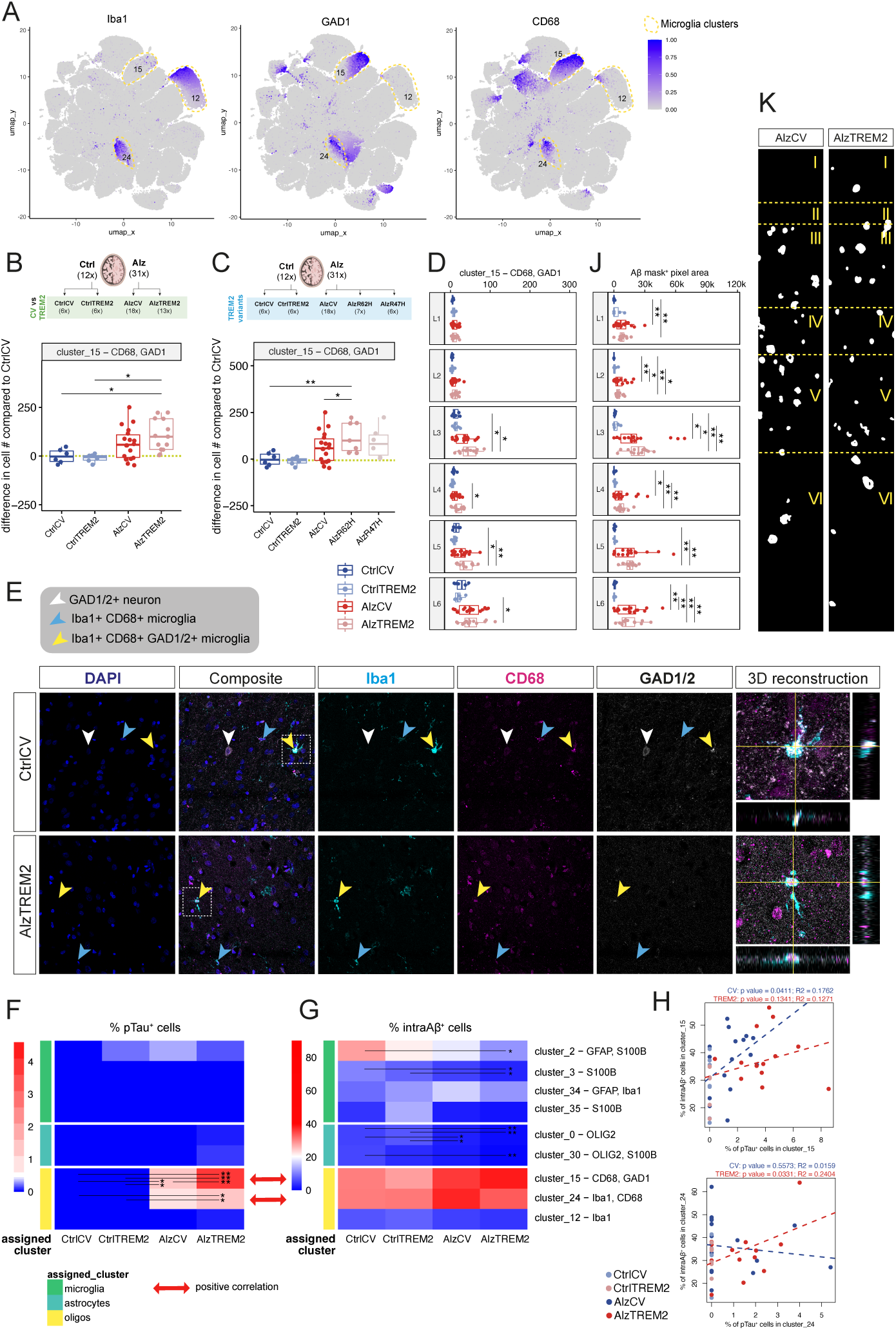
Increased microglia reactivity is associated with markers of vulnerable neurons, pathological proteins and plaques. (**A**) UMAP plots of all clustered nuclei, highlighting expression of the three main markers identified in microglia clusters (Iba1, CD68, GAD1). (**B**,**C**) Difference in average number of CD68^+^GAD1^+^ cells (cluster_15) compared to CtrlCV. Sample were grouped for expression of TREM2 common allele or risk variants, as indicated in the schematic above. Yellow dashed line indicate the Y value of 0. (**D**) Layer-specific increase in CD68^+^GAD1^+^ cells (cluster_15). (**E**) Triple immunostaining for Iba1 (cyan), GAD1 (grey), CD68 (magenta) and DAPI (blue) of CtrlCV and AlzTREM2 samples (left) with 3D reconstruction (right) of the region indicated by the white dashed line, showing triple positive Iba1^+^CD68^+^GAD1^+^cells (yellow arrowhead). (**F**,**G**) Proportion of pTau^+^ (F) and intraAβ^+^ (G) cells in each glial subtype cluster. Coloured boxes on the left highlight groups of glial types (microglia, astrocytes and oligodendrocytes) to which clusters were assigned to (“assigned_cluster”). Red arrows indicate positive correlation shown in (**H**). (**H**) Correlation between pTau^+^ and intraAβ^+^ cells within two CD68+ reactive microglia clusters along disease progression calculated as linear regression (*p* and *R^2^* values indicated above) in CV and TREM2 samples separately (blue and red colour, respectively). (**J**,**K**) Layer-specific quantification of Aβ mask^+^ pixel area (J) generated from the Aβ channel to identify plaques (example of plaques masks from AlzCV and AlzTREM2 samples; K). Statistical significance was calculated with Dirichlet regression (B,C) or ANOVA and Tukey tests or Kruskal–Wallis and Wilcoxon signed-rank test for normally and not normally distributed data respectively (D,F,G,J). Data expressed as percentage (F,G) was transformed with *arcsine* before proceeding with normality and statistical tests.

**Figure 5.**
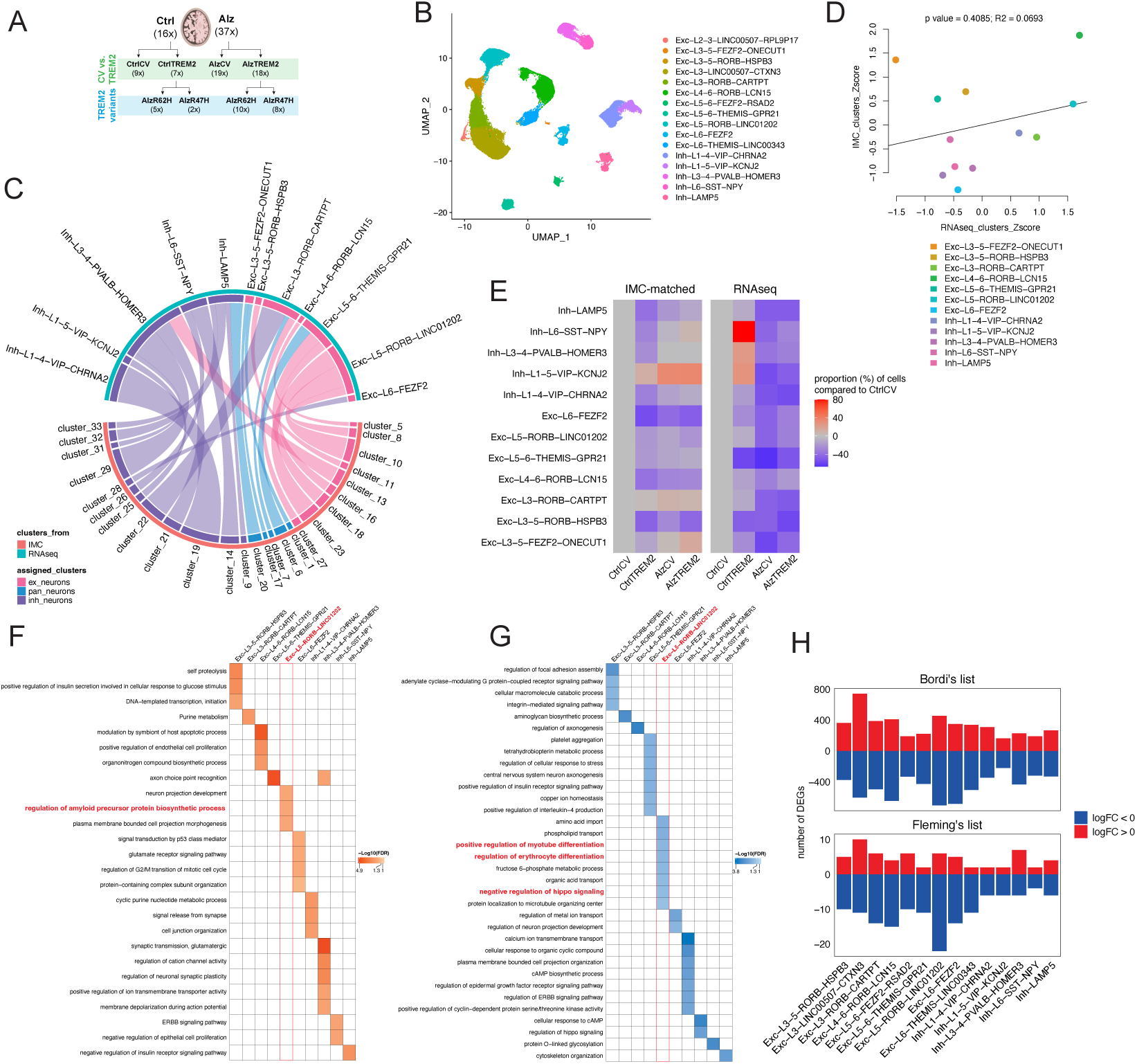
DEGs and pathway analysis of IMC-RNAseq matched neuronal clusters. (**A**) Samples cohort used for snRNAseq analysis in^35^ (43/53 corresponding to the same samples used in this study). (**B**) UMAP plot showing identified clusters of MTG-derived neuronal subpopulations from snRNAseq dataset. (**C**) Final matches of neuronal clusters from IMC (orange, below) and snRNAseq (green, above) experiment. Colour and thinness of the connecting bands identify neuronal types (excitatory, inhibitory and pan-neurons) to which clusters were assigned to (“assigned_cluster”) and matching score, respectively (thicker band implies higher matching score). (**D**) Correlation between Z scores of relative size of matched clusters in IMC (y axis) and RNAseq (x axis), quantified as linear correlation (*p* and *R^2^* values indicated above). (**E**) Proportional increase or decrease in cell number in each RNAseq and matched IMC cluster compared to CtrlCV. (**F**,**G**) Pathway analysis of RNAseq matched clusters regressed for amyloid deposition, showing upregulated (F) and downregulated (G) pathways. Relevant pathway altered in Exc−L5−RORB−LINC01202 cluster are highlighted in red. (**H**) Number of upregulated (red) and downregulated (blue) DEGs from Bordi or Fleming lists in each RNAseq cluster.

### Pre-processing and quality-control of snRNA sequencing data

Alignment and demultiplexing of raw sequencing data was performed using 10X Genomics Cell Ranger v3.1, with a pre-mRNA GRCh38 genome reference^44^ including both introns and exons. Downstream primary analyses of gene-cell matrices were performed using our scFlow pipeline^64^. Ambient RNA profiling was performed using emptyDrops with a lower parameter of <100 counts, an alpha cut-off of ≤0.001, and with 70,000 Monte-Carlo iterations^65^. Cells were filtered for ≥400 and ≤40000 total counts and ≥200 and ≤20000 total expressive features, where expressivity was defined as a minimum of 2 counts in at least 3 cells. The maximum proportion of counts mapping to mitochondrial genes was set to 5%. Doublets were identified using the DoubletFinder algorithm, with a doublets-per-thousand-cells increment of 8 cells (recommended by 10X Genomics), a pK value of 0.005, and embeddings were generated using the first ten principal components calculated from the top 2000 most highly variable genes (HVGs)^66^.

### Integration, clustering, and visualization of data

The linked inference of genomic experimental relationships (LIGER) package was used to calculate integrative factors across samples^67^. LIGER parameters used included: k: 50, lambda: 5.0, thresh: 0.0001, max_iters: 100, knn_k: 20, min_cells: 2, quantiles: 50, nstart: 10, resolution: 1, num_genes: 3000, center: false. Two-dimensional embeddings of the LIGER integrated factors were calculated using the uniform-manifold approximation and projection (UMAP) algorithm with the following parameters: pca_dims: 30, n_neighbours: 50, init: spectral, metric: euclidean, n_epochs: 200, learning_rate: 1, min_dist: 0.4, spread: 0.85, set_op_mix_ratio: 1, local connectivity: 1, repulsion_strength: 1, negative_sample_rate: 5, fast_sgd: false (McInnes et al., 2018). The Leiden community detection algorithm was used to detect clusters of cells from the UMAP (LIGER) embeddings; a resolution parameter of 0.0001 and a k value of 50 was used^68^.

### Assigning cell type labels to snRNAseq cells

Automated cell-typing was performed essentially as previously described using the Expression Weighted Celltype Enrichment (EWCE) algorithm in scFlow against a previously generated cell-type data reference from the Allan Human Brain Atlas^64,69^. The top five marker genes for each automatically annotated cell-type were determined using Monocle 3 and validated against canonical cell-type markers^70^.

### Re-clustering of the neuronal sub-population

Excitatory and inhibitory neuronal clusters were first sub-setted from the single cell object generated in the initial clustering and cell type annotation. Data was first normalized and scaled using Seurat’s NormalizeData and ScaleData functions respectively. RunPCA function was used to calculate the first 20 PCs using the top 2000 highly variable genes. Individual samples were re-integrated with Harmony^80^, using Seurat’s RunHarmony() function (group.by.vars = “manifest”). To produce the final UMAP, we used the following parameters in RunUMAP() (dims = 1:20, n.epochs = 500). To identify clusters, we used first the function FindNeighbors() (dims = 1:20) and then performed unbiased clustering by using FindClusters() (resolution = 0.5). Automated cell-typing was performed essentially as previously described using the Expression Weighted Celltype Enrichment (EWCE) algorithm in scFlow against a previously generated cell-type data reference from the Allan Human Brain Atlas^64,69^

### Dirichlet modelling of relative cell-type composition

To identify changes in relative cell-type composition across categorical variables (e.g., TREM2var vs CV), we used a Dirichlet-multinomial regression^71^ which accounts for dependencies in cell-type proportions within samples while accounting for additional co-variates (e.g., sex + age + APOE + CD33). Threshold for significance was set at an adjusted p-value <0.05.

### Differential gene expression analysis

We used model-based analysis of single-cell transcriptomics (MAST) to identify genes differentially expressed (associated) with histopathological features (using 4G8 amyloid, and PHF1), using each feature as a dependent variable in a zero-inflated regression analysis using a mixed-model^72^. Additionally, diagnosis (control, AD) was used as a dependent variable to identify DGE between experimental groups. Models were fit separately for each cell-type, with and without stratification by TREM2 genotype (none, *TREM2var*, *R47H*, or *R62H*). The model specification was zlm(∼dependent_variable + (1|sample) + cngeneson + pc_mito + sex + age + APOE + CD33, method = “glmer”, ebayes = F). The fixed-effect term cngeneson is the cellular detection rate as previously described, and pc_mito accounts for the relative proportion of counts mapping to mitochondrial genes. Each model was fit with and without the dependent variable and compared using a likelihood ratio test. Genes expressed in at least 10% of cells (minimum of 2 counts per cell) were evaluated for gene expression. The threshold for significant differential gene expression was a log2 fold-change of at least 0.25 and an adjusted p-value < 0.05.

### Impacted pathway analysis

Impacted pathway analysis (IPA) was performed essentially as previously described using the enrichR packages in scFlow^73, 74^. Statistically significant differentially expressed genes were submitted for IPA with the over-representation analysis (ORA) enrichment method against the ‘GO_Biological_Process’, and ‘KEGG’ databases. The false-discovery rate (FDR) was calculated using the Benjamini-Hochberg method and filtering was applied at a significance threshold of ≤0.05. Pathways were selected based on the removal of pathways with FDR > 0.05, single genes, pathways identified with other cells, over-lapping descriptions and non-specific descriptions.

### Matching of IMC and RNA neuronal clusters

For matching IMC and snRNAseq neuronal clusters, all non-neuronal IMC clusters and markers as well as genes highly expressed in all snRNAseq clusters (MAP2, APP, RBFOX3 and SYP; Fig. S6.A) were excluded from scoring calculation. Average expression of neuronal markers used in the IMC experiment was extracted from snRNAseq neuronal clusters (derived from reads number; Fig. S6.A), and average markers expression in both snRNAseq and IMC neuronal clusters were scaled to an equal range (-2 to 5) within each of the markers, separately. The 7 most expressed markers in each snRNAseq and IMC neuronal cluster were identified. The similarity scores for each IMC-snRNAseq clusters combination (Table 4 – all scores; Fig. S6.B) were calculated by summing the scaled expression of the shared top 7 markers (“common_markers”) in the IMC and RNAseq dataset separately (“score_IMC” and “score_RNA”), which were then multiplied together (“combined_score”). The highest similarity score for each IMC-snRNAseq clusters combination was used to identify matching clusters (Table 4 – final matches; Fig. 5.C). The best method for markers scaling range and number of top expressed markers to consider was tested (Fig. S1.B) by checking the proportion of correctly assigned clusters among the most obvious (correct IMC-RNA matches) and by minimizing the number of snRNAseq clusters left unmatched (number of RNA clusters matched out of the total 16; Fig. S1.C-D). Method A was deemed as the most appropriate as it identified the same number of correct matches as second best methods C and D, but leaving less snRNAseq clusters unmatched (only 4, compared to 8 and 7 for methods C and D respectively).

## RESULTS

### Multi-plexed immunohistology identified neuronal subtypes, their cortical organisation and associated glia

We used a 31-antibody panel optimised for IMC (Fig. 1.B) for multiplexed immunostaining of *post-mortem* FFPE MTG sections of 12 non-diseased controls and 31 AD samples (Fig. 1.D). Upon IMC images acquisition and automated processing with SIMPLI^42^, we identified 237,248 cells (1839±277 cells/ROI), of which 198,470 expressed at least one marker. These were clustered (Fig. 1.G) and subsequently assigned to neuronal or glial subpopulations based on markers expression in each cluster (Fig. 1.E-F).

The relative proportions of excitatory (which included pan-neuronal and synapses clusters) and inhibitory neurons detected were as expected^45^, representing 72±2% and 28±2% of total neurons, respectively (Fig. 1.H). The proportion of astrocytes to total glial cells (45±6%) also matched the expected ratio, although the relative abundance of oligodendrocytes (26±6%) and microglia (28±3%) were under- and over-represented^46^, respectively, suggesting limitations in use of a single marker protein for oligodendrocytes at the spatial resolution achieved. Our glial markers identified reactive (cluster_2, cluster_34) and non-reactive (cluster_3, cluster_33) astrocyte phenotypes, as well as those for homeostatic (cluster_12) and activated microglia (cluster_15, cluster_24).

Neuronal subtypes were found to have layer-specific distributions that generally were consistent with those expected (e.g., PCP4^+^ neurons in L5 [cluster_23] and CUX2^+^ neurons in L2/3 [cluster_13]^47^; PVALB^+^ neurons L4/5 [cluster_29] and SST+ L5/6 [cluster_14]^48^) (Fig. 1.J). These results thus provided confidence that the multiplexed IMC could be used to quantify the relative abundance of neuronal subtypes, their cortical localisations and spatial relationships with activated microglia and astrocytes in AD.

### RORB^+^ and GAD1^+^ neurons account for majority of neuronal loss in AD

We first set to determine which cell populations were selectively lost in sporadic AD cases (AlzCV; n=18) relative to non-diseased control donors carrying the common *TREM2* allele (CtrlCV; n=6) (Fig. 2.A). Compared to CtrlCV, the total population of excitatory (-13.5±2%), inhibitory (-11.5±2%) and pan- (-2±0.3%) neurons were not reduced significantly in AD samples (*p*-value from *Dirichlet regression* = 0.3787; Fig. 2.B).

To test whether relative numbers of specific neuronal subtypes decreased with AD, we assessed differences in the numbers of cells identified in distinguishable neuronal sub-type clusters (Fig. 2.C). Compared to CtrlCV, 6 neuronal clusters were significantly reduced in size in AlzCV donors, half of which expressed RORB (RORB^+^MAP2^+^ [cluster_5], RORB^+^FOXP2^+^ [cluster_8], RORB^+^ [cluster_11]). These accounted for 65% of the total neuronal loss (253 RORB^+^ neurons from the total of 388 in clusters that were significantly smaller with AD). The other clusters showing decreased neuronal numbers with AD corresponded to GAD1^+^ADARB1^+^ (cluster_9) and ADARB1^+^ (cluster_28) inhibitory neurons.

We then explored layer-specific localisations for these vulnerable neuronal sub-types in AlzCV samples compared to controls (Fig. 2.D). We found RORB^+^FOXP2^+^ neurons selectively lost in L3,4 and 6 (cluster_8). Vulnerable GAD1^+^ADARB1^+^ neurons were reduced significantly in L4 (cluster_9), although most of these neurons were localised to L6. GAD1^+^FOXP2^+^ inhibitory neurons (cluster_32) were reduced in L3-6. Highlighting the layer specificity of neuronal sub-type vulnerability, RORB^+^GPC5^+^ excitatory neurons (cluster_10) were significantly reduced in number in L1 but relatively increased in number in L6. Together, these results identify L3,4 and 6 RORB^+^ cells as the major neuronal excitatory subtype selectively lost in sporadic AD, consistent with some previous studies^6,7^. We also have defined evidence for layer specific (L1) GAD1^+^ inhibitory neuron loss.

### Neuronal loss is greater amongst donors carrying TREM2 risk variants

To further address whether TREM2 risk variants are associated with increased neuronal loss^35^, we extended our analysis to include samples from non-diseased control (CtrlTREM2; n=6; 2/6 carrying the *R47H* allele) and AD (AlzTREM2; n=13; 6/13 carrying *R47H* allele) heterozygote donors expressing *TREM2 R47H/R62H* variants (Fig. 2.E). Despite small differences compared to AlzCV, the total number of excitatory (+3.6±0.5%), inhibitory (-11.5±2%) or pan- (+7±1%) neurons (*p*-value from *Dirichlet regression =* 0.7738), was not significantly reduced in AlzTREM2 cases (Fig. 2.F,G)

We identified the same neuronal subtype-specific vulnerabilities in AlzTREM2 samples relative to CtrlCV that we found for the AlzCV with lower numbers of RORB^+^MAP2^+^ (cluster_5), RORB^+^FOXP2^+^ (cluster_8), RORB^+^ (cluster_11), GAD1^+^ADARB1^+^ (cluster_9) and ADARB1^+^ (cluster_28) neuronal sub-types (Fig. 2.G). However, the relative loss in AlzTREM2 samples was greater than for AlzCV (36.3±15.8% RORB^+^FOXP2^+^ [cluster_8], 27.6±15.9% RORB^+^ [cluster_11] and 16.1±13.7% ADARB1^+^ [cluster_28] less cells in AlzTREM2 compared to AlzCV cases). A GAD1^+^ cluster also was significantly reduced in AlzTREM2 samples compared to CtrlCV (GAD1^+^FOXP2^+^ [cluster_32]: -25.9±20.7%). However, we found relatively more RORB^+^GPC5^+^ neurons (cluster_10) in AlzTREM2 compared to CtrlTREM2 (+19.8±12.5%, compared to AlzCV).

Neurons also were lost preferentially in specific layers: L5,6 RORB^+^FOXP2^+^ neurons (cluster_8), L6 GAD1^+^FOXP2^+^ neurons (cluster_32) and L5 GAD1^+^ neurons (cluster_33; Fig. 2.H). GAD1^+^ADARB1^+^ neurons (cluster_9), although most abundant in L6, show a small but significant reduction in L1 (AlzCV vs. CtrlTREM2; *p*-value from *Kruskal Wallis test* = 0.024). The greatest relative increase in numbers of RORB^+^GPC5^+^ neurons (cluster_10) was found in L3.

These results indicate that expression of TREM2 risk variants enhances loss predominantly for the same neuronal sub-types most vulnerable in AlzCV cases. L3 RORB^+^GPC5^+^ neurons appear to be relatively resilient.

### TREM2 R47H risk variant is associated with greatest neuronal loss in AD

To explore which TREM2 risk variant was associated with the greatest neuronal loss in AD, we re-analysed our dataset splitting AlzTREM2 samples based on their *TREM2* genotype (AlzR62H [n=7] and AlzR47H [n=6]; Fig. 2.J). Among the 6 neuronal subpopulations showing evidence for selective loss in AlzTREM2 cases, 4 were significantly reduced in AlzR47H cases compared to CtrlCV (RORB^+^MAP2^+^ [cluster_5], RORB^+^ [cluster_11], GAD1^+^ADARB1^+^ [cluster_9] and ADARB1^+^ [cluster_28]; Fig. 2.L). RORB^+^FOXP2^+^ (cluster_8) neurons were significantly reduced in both TREM2 variants compared to CtrlCV, while the NTNG2^+^LMO3^+^ cluster_20 had relatively increased numbers of neurons only in AlzR62H samples compared to AlzCV. AlzR62H samples show relative loss of RORB^+^FOXP2^+^ (cluster_8) cells in L2 and AlzR47H samples showed PCP4+ (cluster_23) cells in L6 (Fig. 2.M). Together, these results suggest that, although relatively greater numbers of neurons appear to be lost, the same sub-types of neurons are selectively vulnerable in AD associated with *R47H TREM2* and CV AD.

### Neuronal subtypes accumulating NFT are relatively resilient

To explore the relationships between ptau accumulation to neuronal vulnerability, We quantified relative amounts of extra- and intra-and cellular ptau. For this analysis, AlzCV samples were split into early- (Braak 3-4) and late-AD (Braak 5-6) cases (Fig. S4.B). As expected, we found the total ptau immunostaining (quantified as pixel area; Fig. S4.A) increases with Braak stage compared to non-diseased controls and earlier AD stages (Fig. S4.C). A shift towards higher extra-cellular ptau signal is visible in AlzTREM2 cases (Fig. S4.C), with no apparent differences between TREM2 genotypes (Fig. S4.F,G). Also as expected, intra-cellular ptau (NFT) was more frequently found inside neurons in brains with higher Braak stage (Fig. S4.D). The proportion of NFT^+^ cells was positively correlated with extra-cellular deposition of both Aβ (*R^2^* = 0.40) and pTau (*R^2^* =0.86) in CV and *TREM2* variant (Aβ, *R^2^* = 0.77; pTau, *R^2^* = 0.78) (Fig. S4.E).

To test whether NFT accumulate preferentially in any specific neuronal subpopulation, we quantified the proportion of NFT^+^ cells in each cluster. This was highest among RORB^+^GPC5^+^ (cluster_10; 46.4±32.3%) neurons, for which the relative numbers of ptau^+^ neurons increased with disease progression (Fig. 3.A,C). A lower proportion was found in the NPY^+^ (cluster_26; 2.4±3.5%), GAD1^+^FOXP2^+^ (cluster_32; 1±5.1%) and Calretinin^+^ (cluster_19; 1±1.5%) neuronal sub-types, but did not significantly increase with disease progression. RORB^+^pTau^+^ cells were found in AlzCV samples mostly in regions corresponding to L3 (Fig. 3.D), where RORB^+^GPC5^+^ (cluster_10) neurons were identified to be increasing in relative numbers with disease progression (Fig. 2.H). These results together suggest that ptau is a marker of relative neuronal resilience, rather than vulnerability.

### Intraneuronal accumulation of Aβ is a marker of vulnerability to cell death

We then tested for correlations between extra-cellular Aβ and neuronal loss. Total Aβ^+^ area (Fig. S4.A) increased in later relative to early Braak stages or non-diseased controls (Fig. S4.C). AlzTREM2 cases had greater extra-cellular Aβ deposition compared to AlzCV, however not significant (Fig. S4.C).

We assessed the proportion of intraAβ^+^ cells in each neuronal sub-type cluster. Overall, the proportion of intraAβ^+^ cells in neuronal cluster was highest in CtrlCV (23.4±6.4%), lower in early Braak 3-4 AlzCV (16.8±7.6%; *p* value = 0.463) and lowest in late Braak 5-6 AlzCV (14.4±4.9%; *p* value = 0.031) and AlzTREM2 samples (10.2±4.7%; *p* value = 0.000; Fig. S4.D). The proportion of intraAβ^+^ cells was negatively correlated with both extra-cellular Aβ (CtrlCV-AlzCV*, R^2^* = 0.20; CtrlTREM2-AlzTREM2, *R^2^* = 0.52) and pTau (CtrlCV-AlzCV*, R^2^* = 0.08; CtrlTREM2-AlzTREM2 *R^2^* = 0.59) (Fig. S4.E). These results suggest accumulation of intraAβ early with progression of pathology.

The proportion with intraAβ^+^ was highest in RORB^+^FOXP2^+^ (cluster_8; 99.2±1.8%) and GAD1^+^FOXP2^+^ (cluster_32; 99.7±1.9%) neurons (Fig. 3.B,C). A lower proportion was also found in CALB1^+^ (cluster_22; 32.19±17.1%), Calretinin^+^ (cluster_19; 28,3±15.6) and PVALB^+^CCK^+^ (cluster_29; 25±15.5%) neurons (Fig. 3.B). Apart from RORB^+^FOXP2^+^ (cluster_8) and GAD1^+^FOXP2^+^ (cluster_32) neurons, the proportion of intraAβ^+^ cells within these clusters was lower with greater Braak stages. IntraAβ^+^ RORB+ and GAD1+ neurons were found predominantly in regions corresponding to L3-6 (Fig. 3.E,F) in which the majority of RORB^+^FOXP2^+^ (cluster_8) and GAD1^+^FOXP2^+^ (cluster_32) cells had previously been localised (Fig. 2.D,H).

Our results thus show that intraneuronal Aβ and pTau preferentially accumulate in different neuronal subtypes (intraAβ in RORB^+^FOXP2^+^ and GAD1^+^FOXP2^+^; pTau in RORB^+^GPC5^+^). The lower number of intraAβ^+^ neurons with higher Braak stage suggests that the neuronal sub-types expressing intraAβ^+^ are selectively lost early in the progression of pathology; intraAβ accumulation appears to be a marker of vulnerability to neuronal death. By contrast, pTau is localised preferentially in a neuronal sub-types that appear to be relatively resilient to neuronal loss.

### Phagocytic microglia and β-amyloid plaques are spatially associated with vulnerable neurons

Microglia phagocytose synapses in the context of pathological β-amyloid in AD. We identified that clusters of reactive glia including a marker of inhibitory synapses marker (CD68^+^GAD1^+^ [cluster_15], Fig. 4.A,B) was more abundant with AD than in non-disease control tissues. There also was a trend to greater abundance of Iba1^+^CD68^+^ [cluster_24] microglia (Fig. S5.B). Greater increases in numbers of reactive microglia were found in tissues expressing *TREM2* risk variants, particularly *R62H* (Fig. 4.C). Reactive astrocytes (GFAP^+^S100β^+^ [cluster_2]; Fig. S5.A) also were more abundant with AD (Fig. S5.C) and particularly in tissues expressing *TREM2* risk variants (Fig. S5.D).

Activated glia also appeared to phagocytose ptau and β-amyloid; reactive CD68^+^ microglial clusters (cluster_15, cluster_24) showed the highest proportions of intracellular ptau and Aβ (Fig. 4.F,G). The relative proportions of microglia with intracellular ptau and Aβ was greater in tissues showing greater disease progression. Intracellular pTau^+^ cells was found particularly amongst CD68^+^GAD1^+^ (cluster_15) and Iba1^+^CD68^+^ (cluster_24) reactive microglia in AlzTREM2 cases compared to controls. The proportions of CD68^+^ microglia with intracellular ptau and Aβ were correlated and greater with higher Braak stage (CtrlCV-AlzCV in CD68^+^GAD1^+^ [cluster_15] *R^2^* = 0.1762; CtrlCTREM2-AlzTREM2 in Iba1^+^CD68^+^ [cluster_24] *R^2^* = 0.1762; Fig. 4.H [and also see red arrows in Fig. 4.F,G]). Intracellular Aβ^+^ also was detected in astrocytes and oligodendroglia but in lower proportions. Their relative proportions were lower with greater tissue pathological progression.

We validated the co-localisation of GAD1, CD68 and Iba1 in microglia in AD with immunofluorescence. In both control and AlzTREM2 cases, we identified Iba1^+^CD68^+^ cells with evidence of GAD1 signal (yellow arrowheads in Fig. 4.E), which the 3D reconstruction confirmed to be located intra-cellularly. These results together suggest that activated microglia involved in the clearance of pathological ptau and β-amyloid also contribute to the pathological pruning of inhibitory synapses and consequent neuronal hyperexcitability. They also provide some evidence that activated astrocytes may play a secondary role in pathological protein clearance more prominent in earlier stages of disease.

We identified spatial clustering between activated microglia with cortical layers in which vulnerable neurons are preferentially found. We observed a higher abundance of CD68^+^GAD1^+^ neurons in L3, 5 and 6 in AlzTREM2 cases compared to controls Fig. 4.D). A similar but non-significant trend also was apparent in AlzCV samples. By contrast, reactive astrocytes were mostly localised in L1 and 6 and did not show any layer-specific AD-related activation (Fig. S5.E).

To test whether activated microglia and astrocytes were more common near β-amyloid plaques, we quantified the number of reactive GFAP^+^S100B^+^ astrocytes (cluster_2) and CD68^+^GAD1^+^ microglia (cluster_15; Fig. S5.J,K red and blue arrowheads, respectively) within or around the amyloid (indicated as “plaque mask” and “50μm ring”, respectively). Numbers of CD68^+^GAD1^+^ reactive microglia were significantly higher both within and around plaques (Fig. S5.H). Reactive astrocytes were more weakly associated with plaques (Fig. S5.G).

Finally, we analysed whether β-amyloid deposition (and the associated activated microglia) was layer-specific. We identified the greatest relative β-amyloid plaque area (quantified as pixel area of their mask) increase in all cortical layers, with no differences between AlzTREM2 and AlzCV (Fig. 4.J,K). This was particularly evident in L3 (29.4% of total plaque area), L5 (20.8% of total plaque area) and L6 (20.6% of total plaque area) in AlzCV and AlzTREM2 tissues. Thus, β-amyloid plaques densities are greatest in cortical layers also shower higher microglial activation and proportions of vulnerable neuronal sub-types (Fig. 4.D).

### Exploration of vulnerable neuronal phenotypes with paired snRNAseq and multiplexed immunohistology

To explore pathways and molecular mechanisms which may underly vulnerability of the neuronal subpopulations identified, we analysed a snRNAseq dataset generated from the same sample cohort with the addition of 10 more samples to increase the power for differential gene expression analyses^35^ (3 CtrlCV, 1 CtrlTREM2, 1 AlzCV, 5 AlzTREM2; Fig. 5.A; Table 1). snRNAseq data from MTG nuclei were clustered to distinguish those from transcriptomic sub-types excitatory and inhibitory neurons (Fig. 5.B). These transcriptomic were then matched to neuronal sub-types identified by IMC (Fig. 5.C) based on the scores for shared marker expression and abundance (snRNAseq) or intensities (IMC) (Fig. S6.A,B; Table 4).

The majority of sub-types of excitatory (8/9) and inhibitory (10/12) neurons defined immunohistologically by IMC clusters were able to be matched with a corresponding transcriptomic excitatory and inhibitory sub-type. Pan-neuronal IMC clusters almost exclusively (4/5) matched with RNAseq excitatory neuronal clusters (Fig. 5.C). Most matched clusters showed similar relative sizes (Z-score [Fig. 5.D], proportion of cells/nuclei per cluster [Fig. S6.C]). Relative differences in sizes of clusters between types of samples (AD or non-diseased controls) also were similar between IMC and RNAseq matched clusters (Fig. 5.E; Fig. S6.D). Both transcriptomically-defined Exc−L5−RORB−LINC01202 and IMC RORB^+^FOXP2^+^ (cluster_8) neurons were significantly reduced in size in AD cases compared to controls (Fig. S6.E).

### APP biosynthesis and autophagy pathways are differentially expressed with AD pathology in vulnerable RORB^+^ neuronal clusters

To explore molecular mechanisms potentially underly neuronal vulnerability, we performed differential gene expression (DEGs; Fig. 5.H: Fig. S.8,9; Table 6) and pathway (Fig. 5.F,G; Fig. S.7; Table 5) analyses of RORB^+^-associated snRNAseq neuronal clusters separately contrasted for diagnosis (AD or non-diseased control) or by regressing for tissue PHF1^+^ ptau or 4G8^+^ amyloid denisities^35^.

Genes and pathways differentially expressed with greater amyloid deposition in the Exc−L5−RORB−LINC01202 cluster included upregulation of genes known to be involved in pathological Aβ fragment production (*ABCA2*^49^, *NECAB3*^50^ also called *XB51/NIP1,* and *ITM2B*^51^ from the “regulation of amyloid precursor protein biosynthetic process” pathway; Fig. 5.F). Mitogen-activated protein kinase 14 (*MAPK14*), a deficiency or inhibition of which potentiates lysosomal activity for degradation of Aβ^52^, was downregulated (from “positive regulation of myotube differentiation”, “regulation of erythrocyte differentiation” and “negative regulation of hippo signalling” pathways; Fig. 5.G). We also found pathways related to protein misfolding, such as “response to unfolded protein” in Exc-L4-6-RORB-LCN15, Exc-L6-FEZF2 and Inh-L3-4-PVALB-HOMER3 clusters and “chaperone cofactor-dependent protein refolding” in Exc-L5-RORB-LINC01202 downregulated in AD cases relative to non-diseased controls (Fig. S7.B). Amongst genes differentially reduced in expression with AD was mortalin (*HSPA9*), a heat shock mitochondrial protein previously associated to degenerative neurons reactivity to stress in Parkinson’s disease^53^. Moreover, downregulation of *HSPA1A*, which encodes HSP70, a heat shock protein which inhibits aggregation of Aβ^54^, was found in all of the neuronal clusters. Thus, while abnormal APP processing and lysosomal genes were specifically dysregulated with AD in the vulnerable Exc−L5−RORB−LINC01202 neurons, a transcriptomic phenotype consistent with impaired protein folding mechanisms was expressed by most neuronal subpopulations.

To explore the relative expression of genes of the autophagy-lysosomal pathway previously associated with AD pathology including intraAβ accumulation in amyloid mouse models^19,20^, we tested for differential expression of genes involved in autophagy^55^ and neurodegenerative diseases (AD and FTD; Table 6)^56^. We found that majority of genes tested were downregulated relatively specifically in the Exc−L5−RORB−LINC01202 cluster (Fig. 5.H, S9, 10). Indeed, the Exc−L5−RORB−LINC01202 cluster matches with both RORB^+^FOXP2^+^ (cluster_8) and RORB^+^GPC5^+^ (cluster_10) cells, which show the highest accumulation of intraAβ and pTau, respectively (Fig. 3.A,B). This provides evidence that defective APP biosynthesis and autophagy-lysosomal processes contribute to the vulnerability of Exc−L5−RORB−LINC01202 neurons (matched to both RORB^+^FOXP2^+^ and RORB^+^GPC5^+^ neuronal sub-types) with greater AD pathology.

## DISCUSSION

We combined IMC with snRNAseq to identify the first inhibitory and excitatory neuronal subpopulations lost in early AD in the MTG, and the intrinsic characteristics underlying their vulnerability. We show evidence of selective loss among L2,3 and 5,6 RORB^+^ and L3,5 and 6 GAD1^+^ neurons with AD. These subpopulations also prominently accumulate intraAβ, while pTau progressively increases in a spatially distinct neuronal subtype, suggesting intraAβ, rather than pTau, to represent the initial pathological event leading to selective neuronal loss in AD. We show that both astrocytes and microglia are more reactive in AD samples, but microglia is more strongly associated with clearance of both vulnerable neurons and pathological proteins. Expression of TREM2 *R47H/R62H* variants leads to a variant-specific increase of both neuronal loss and glial reactivity, underlying their more severe pathology. Finally, transcriptomic analyses identifies a neuronal subtype-specific dysregulation of autophagy-lysosomal genes which may predispose to accumulation of pathological proteins and higher vulnerability. Our study describes how intraAβ accumulation, and the potentially underlying molecular mechanisms, may be linked to layer- and neuronal subtype-specific vulnerability to cell death, representing the earliest pathological events in human AD.

Majority of vulnerable neurons markers identified in our study were previously established in other brain regions (RORB^+^ and GAD1^+^ in EC L2^7^ and PFC^47^, respectively) however using different methodologies (snRNAseq). This both validate our findings and points towards a potential common mechanism underlying neuronal vulnerability. A subset of vulnerable neurons identified also show expression of FOXP2, a marker recently associated to AD vulnerable neurons too^57^. Conversely, ADARB1^+^ neurons are our only population not previously identified among AD vulnerable neurons. However, reduction in ADARB1 expression is associated with AD^58^. On the other hand, we do not find a significant reduction in total number of other neuronal subpopulations previously identified as vulnerable in AD, such as CUX2^+^, PCP4^+^, GPC5^+^ and SST^+^ neurons^7,35,47^. Nevertheless, a tendency towards layer-specific loss is visible for PCP4^+^, GPC5^+^ and SST^+^ subtypes, particularly in L6, suggesting our analysis might be limited by a reduced number of detected cells.

We show for the first time in the human brain that intraAβ^+^ cells progressively reduce from controls to higher Braak stages, previously only observed in AD mouse models^26^. This suggests intraAβ burdened cells either can clear intraAβ or are lost due to cell death. The identification of both selectively lost neuronal populations as the major intraAβ carrier strongly suggest the second hypothesis. Indeed, not all selectively lost neuronal populations show high intraAβ signal (e.g., ADARB1^+^ neurons) and *vice versa*, not all neuronal populations showing fairly higher intraAβ signal are reduced in number (e.g., Calretinin, CALB1, PVALB, PCP4). Calretinin^+^, CALB1^+^ and PVALB^+^ inhibitory neurons are known resilient populations in AD^1^ and this decoupling might further underly their resistance. Conversely, the mild non-significant reduction in PCP4^+^ cells in L6 suggests intraAβ-lead death might occur in this neuronal subtype too. Specific NFT formation in RORB^+^GPC5^+^ neurons has been recently identified by an independent group^57^, however, in our study this populations is not selectively lost. Our result is in agreement with previous research showing that neuronal loss exceeds the number of NFTs in the MTG^59^, and occurs in NFT-free neurons in the PFC^47^. Nevertheless, NFT-lead neuronal death occurs, as demonstrated by “ghost tangles” in AD *post-mortem* brains^60^. independent or semi-independent mechanism might therefore drive selective neuronal loss.

By including AD patients carrying TREM2 risk variants, we further explored the relationship between neuronal loss, glial reactivity and accumulation of pathological proteins in AD. As previous studies have shown^35^, AlzTREM2 cases are characterized by higher neuronal loss. In our analysis, AlzTREM2 cases also show a lower proportion of intraAβ^+^ cells, which further suggest a causal link between these pathological events. As expected, microglia reactivity is generally higher in donors carrying the TREM2 risk variant compared the common allele, particularly in those carrying *R62H*, as *R47H* is the most detrimental to their reactivity^61^. We do not observe a higher astrocytic reactivity in AlzR47H samples compared to AlzR62H, which was expected as, in mice, astrocytes increase their phagocytosis and GFAP expression for compensating for microglia dysfunctions^62^. Our results also support previous findings^63,64^ suggesting microglia to be the major responsible for clearance of both vulnerable neurons and AD pathological proteins. However, whether intraAβ accumulation in vulnerable neurons triggers microglia reactivity, or *vice versa*, still needs to be determined. A potential cross-talk has been observed in AD mice models, and intraAβ^+^ neurons proposed as the primary pro-inflammatory agent at the pre-plaque stage^33^. Additionally, the lower proportion of intraAβ^+^ cells and higher extra-cellular Aβ deposition observed in AlzTREM2 cases, supports previous theories suggesting amyloid plaques originate from death of intraAβ-burdened neurons^19,65^. In agreement, our results show that plaques, selective loss of intraAβ-burdened neurons and higher microglia reactivity are located in the same cortical layers. This potentially damaging interaction in early AD may lead to identification of new cellular targets and pathological mechanism to prevent neuronal loss.

Our transcriptomic analysis suggests one such pathological mechanism may be impairment of autophagy-lysosomal activity in vulnerable neurons which may lead to intraAβ accumulation, as previously observed in AD mouse models^19,20^. Changes in the autophagy lysosomal pathway specific to AT8^+^ neurons have been observed before in patients affected by AD^47^ and tauopathies^66^, showing however an increase in p62/SQSTM1 and LC3 expression, respectively. We do not observe the same reduction in the expression of autophagy genes in GAD1^+^ vulnerable neurons, however this might be due to the lower granularity of inhibitory neurons populations identified in the snRNAseq compared to IMC. The therapeutic potential of autophagy enhancers, which reduce neuronal loss and cognitive decline in AD mouse models, has long been explored in AD clinical trials^21^. Further validations and investigations of our exploratory analysis will indicate whether targeted enhancement of autophagy activity in vulnerable neurons might support their intraAβ clearance and increase their resilience towards the disease.

Our study shows few discrepancies with previous investigations which might arise from methodological differences. Firstly, classification of neuronal subtypes using mRNA sequencing and antibody-based staining might differ as often mRNA-protein correlations poorly match. Secondly, sensibility and specificity of primary antibodies might be less consistent than mRNA sequencing, particularly for *post-mortem* human brain FFPE sections as processing protocols are high variable among brain banks. Thirdly, a more extensive set of antibodies will be required for grasping the full spectrum of glial subtypes (e.g., ALDH1L1^67^, SOX9^68^) and AD-related activity states (e.g., Aquaporin^69^, NF1A^70^). Similarly, the intra-cellular accumulation of pathological proteins should be further explored using antibodies specific to Aβ-42 (e.g., MOAB-2^71^) and alternative Tau post-translational modifications (e.g., PHF-1 and Ab39^60^) to identify neurons bearing mature tangles. However, despite our neuronal antibody set was broader, some results were unexpected, such as absence of MAP2 and NeuN expression in all neuronal populations, or little co-expression between GAD1 and other inhibitory neuronal markers. Finally, defining neuronal loss as size reduction of a marker-defined cluster relies on each neuronal subpopulation not changing their expression profile with disease progression, which is difficult to prove. Nevertheless, the congruence of our results with previous studies confirm the overall robustness of our approach. Also, our study highlights the benefits of using methods that combine single cells high-throughput transcriptomic data with antibody-based staining, laying the ground for future investigations where both cellular subtype classification and visualisation of pathological proteins are necessary.

In conclusion, our study identifies intraAβ as a marker of selective neuronal vulnerability. Validation of the causative link between these events will define the earliest pathological signs of AD. Whether neuronal subtype specific dysfunction in the autophagy-lysosomal pathway underly intraAβ accumulation needs to be verified but might represent a functional approach for increasing neuronal resilience and delay cognitive impairment in AD. Further analyses will clarify how and when neuronal vulnerability is triggered, whether intraAβ accumulation and defective autophagy are secondary effects due to neuronal morphology and functionality, or intrinsic characteristic ingrained by the molecular mechanisms regulating their embryonic development, as previously suggested^72^.

## Data Availability

All data produced in the present study are available upon request to the authors.

## ACKNOWLEDGEMENTS

We thank the donors and their families for the use of human brain tissue in this study and the UK brain bank staff for making it available. Tissue samples were provided by the London Neurodegenerative Diseases Brain Bank (King’s College London), Newcastle Brain Tissue Resource, Queen’s Square Brain Bank (University College London), Manchester Brain Bank, Oxford Brain Bank, South West Dementia Brain Bank (University of Bristol) and Parkinson’s UK Brain Bank (Imperial College London).

We are grateful to Dr Diana Benitez for her support in the human tissue ordering and management, Dr Michele Bortolomeazzi for his help with SIMPLI troubleshooting and Alan Murphy for his help in data analysis in R studio. PMM acknowledges generous personal support from the Edmond J Safra Foundation and Lily Safra and an NIHR Senior Investigator Award. This work was supported by the UK Dementia Research Institute, which receives its funding from UK DRI Ltd., funded by the UK Medical Research Council, Alzheimer’s Society, and Alzheimer’s Research UK. AC salary and consumable were supported by the UK DRI Cross-centre post-doc programme.

## DECLARATIONS

This study was partly funded by Biogen IDEC. PMM has received consultancy fees from Sudo Biosciences, Ipsen Biopharm Ltd., Rejuveron Therapeutics and Biogen. He has received honoraria or speakers’ fees from Novartis and Biogen and has received research or educational funds from BMS, Biogen, Novartis and GlaxoSmithKline.

---

**Figure S1 – Optimisation of markers threshold and automated IMC-snRNAseq clusters assignment.**

**(A)** Total number of marker^+^ cells obtained applying different thresholds (th) to identified cells (red to blue bars), compared to manual counting (black bars), in one CtrlCV ROI. Optimal threshold for each channel was identified as that yielding the number of marker^+^ cells more similar to manual counting. (**B,C,D**) Optimisation of IMC-snRNAseq clusters matching method. Six different combinations of markers scaling range and number of highest expressed markers to consider (B) were tested by checking assignment of the 7 most of obvious IMC-snRNAseq clusters match (D). Proportion of correct matches (out of 7) and total RNA clusters matched to IMC clusters (out of 16) was used to rate successfulness of each method (C).

**Figure S2 – Cortical layer distribution of all clusters identified.**

Cortical distribution of nuclei within each of the clusters identified (Fig. 1.E). Statistical significance between indicated groups was calculated with ANOVA and Tukey tests or Kruskal–Wallis and Wilcoxon signed-rank test for normally and not normally distributed data respectively.

**Figure S3 – Markers expression in clusters.**

UMAP plots of all clustered nuclei (Fig. 1.G), highlighting expression of all markers used (Fig. 1.B).

**Figure S4 – Total extra-cellular pTau and Aβ accumulation and its correlation to disease progression.**

**(A)** Example IMC images showing total pTau and Aβ signal, quantified in (C,G). (**B,F**) samples grouping based on Braak stage (B) and separate TREM2 variants (F) used for analyses shown in (C,D) and (G, H), respectively. (**C**,**G**) Quantification of total Aβ and pTau signal as ROIs marker^+^ pixel area. (**D**,**H**) Proportion of cells positive for Aβ and pTau signal, representing intraAβ^+^ and NFT^+^ cells, respectively. (**E**) Linear regression showing correlation between proportion of NFT^+^ (left) or intraAβ^+^ (right) cells to total pTau and Aβ pixel area (*p* and *R^2^* values indicated above) in CV and TREM2 samples separately (blue and red colour, respectively).

**Figure S5 – Astrocytes reactivity and glial association to plaques.**

**(A)** UMAP plots of all clustered nuclei, highlighting expression of markers identifying astrocytes (S100B, GFAP) and oligodendrocytes (OLIG2) clusters. (**B**,**C,D**) Difference in average number of GFAP^+^S100B^+^ (cluster_2) and Iba1^+^CD68^+^ (cluster_24) cells compared to CtrlCV. Sample were grouped for expression of TREM2 common allele or risk variants (split by *R62H*/*R47H* in C). Yellow dashed line indicate the Y value of 0. (**E**) Layer-specific changes in number of GFAP^+^S100B^+^ cells (cluster_2). (**F**) Quantification of PLP1 expression as marker^+^ pixel area. (**G**-**K**) Quantification of GFAP^+^S100B^+^ astrocytes (cluster_2; red arrowheads in J) and GAD1^+^S100B^+^ microglia (cluster_15; blue arrowheads in K) within and around plaques (G and H respectively). Mask of plaque area and surrounding 50µm ring were generated with an ImageJ script (example of regions obtained shown in G, J). Statistical significance was calculated with Dirichlet regression (B,C,E) or ANOVA and Tukey tests or Kruskal–Wallis and Wilcoxon signed-rank test for normally and not normally distributed data respectively (D,F,H).

**Figure S6 – Comparison of matched clusters changes in proportional size**

**(A)** Average and proportional expression of markers used in IMC among all snRNAseq neuronal clusters. (**B**) Matching score calculated for all IMC-RNAseq clusters combination. Coloured boxes on the left highlight groups of neuronal types (excitatory, inhibitory and pan-neurons) to which clusters were assigned to (“assigned_cluster”). (**C**) Proportional size of matched clusters in IMC (blue) and snRNAseq (yellow) experiments. (**D**) Correlation between proportional changes in IMC-snRNAseq matched clusters along disease progression, calculated as linear regression (*p* and *R^2^* values indicated for each regression). (**E**) Changes in relative proportion of size of snRNAseq clusters in AD vs. control samples, either carrying TREM2 common allele (above) or risk variants (below) calculated with Dirichlet regression.

**Figure S7 – Pathway analysis of transcriptomic changes in snRNAseq clusters regressed for diagnosis and PHF1 expression**

(**A**-**D**) Upregulated (A,C) and downregulated (B,D) pathways in snRNAseq clusters regressed for diagnosis (A,B) and PHF1 expression (C,D). Altered pathways relevant to in Exc−L5−RORB−LINC01202 cluster or generally to AD are highlighted in red.

**Figure S8 – Analysis of DEGs from Bordi’s list**

(**A**-**J**) Average number of total (A,B), downregulated (D,E) and upregulated (G,H) DEGs within the Bordi’s list per cluster and frequency of the 10 top total (C), downregulated (F) and upregulated (J) DEGs in Exc−L5−RORB−LINC01202 cluster.

**Figure S9 – Analysis of DEGs from Fleming’s list**

(**A**-**J**) Average number of total (A,B), downregulated (D,E) and upregulated (G,H) DEGs within the Fleming’s list per cluster and frequency of the top total (C), downregulated (F) and upregulated (J) DEGs in Exc−L5−RORB−LINC01202 cluster.

## REFERENCES

1 Fu, H., Hardy, J. & Duff, K. E. Selective vulnerability in neurodegenerative diseases. Nature Neuroscience 21, 1350–1358, doi:10.1038/s41593-018-0221-2 (2018).

2 Chin, J. et al. Reelin Depletion in the Entorhinal Cortex of Human Amyloid Precursor Protein Transgenic Mice and Humans with Alzheimer&#039;s Disease. The Journal of Neuroscience 27, 2727, doi:10.1523/JNEUROSCI.3758-06.2007 (2007).

3 Morrison, J. H. et al. A monoclonal antibody to non-phosphorylated neurofilament protein marks the vulnerable cortical neurons in Alzheimer’s disease. Brain Research 416, 331–336, 10.1016/0006-8993(87)90914-0 (1987).

4 Saiz-Sanchez, D., De la Rosa-Prieto, C., Ubeda-Banon, I. & Martinez-Marcos, A. Interneurons, tau and amyloid-β in the piriform cortex in Alzheimer’s disease. Brain Structure and Function 220, 2011–2025, doi:10.1007/s00429-014-0771-3 (2015).

5 Xu, Y., Zhao, M., Han, Y. & Zhang, H. GABAergic Inhibitory Interneuron Deficits in Alzheimer’s Disease: Implications for Treatment. Frontiers in Neuroscience 14 (2020).

6 Otero-Garcia, M. et al. Single-soma transcriptomics of tangle-bearing neurons in Alzheimer’s disease reveals the signatures of tau-associated synaptic dysfunction. bioRxiv, 2020.2005.2011.088591, doi:10.1101/2020.05.11.088591 (2020).

7 Leng, K. et al. Molecular characterization of selectively vulnerable neurons in Alzheimer’s disease. Nature Neuroscience 24, 276–287, doi:10.1038/s41593-020-00764-7 (2021).

8 McInnes, J. et al. Synaptogyrin-3 Mediates Presynaptic Dysfunction Induced by Tau. Neuron 97, 823–835.e828, 10.1016/j.neuron.2018.01.022 (2018).

9 Takashima, A. Mechanism of neurodegeneration through tau and therapy for Alzheimer’s disease. Journal of Sport and Health Science 5, 391–392, 10.1016/j.jshs.2016.08.009 (2016).

10 Dong, Y. et al. Hyperphosphorylated tau mediates neuronal death by inducing necroptosis and inflammation in Alzheimer’s disease. Journal of Neuroinflammation 19, 205, doi:10.1186/s12974-022-02567-y (2022).

11 Kuchibhotla, K. V. et al. Neurofibrillary tangle-bearing neurons are functionally integrated in cortical circuits in vivo. Proceedings of the National Academy of Sciences 111, 510–514, doi:10.1073/pnas.1318807111 (2014).

12 Li, H.-L. et al. Phosphorylation of tau antagonizes apoptosis by stabilizing β-catenin, a mechanism involved in Alzheimer’s neurodegeneration. Proceedings of the National Academy of Sciences 104, 3591–3596, doi:10.1073/pnas.0609303104 (2007).

13 Wu, M. et al. Friend or foe: role of pathological tau in neuronal death. Molecular Psychiatry, doi:10.1038/s41380-023-02024-z (2023).

14 Cras, P. et al. Senile plaque neurites in Alzheimer disease accumulate amyloid precursor protein. Proc Natl Acad Sci U S A 88(17), 7552–7556, doi:10.1073/pnas.88.17.7552 (1991).

15 LaFerla, F. M., Green, K. N. & Oddo, S. Intracellular amyloid-β in Alzheimer’s disease. Nature Reviews Neuroscience 8, 499–509, doi:10.1038/nrn2168 (2007).

16 Ripoli, C. et al. Intracellular Accumulation of Amyloid-β (Aβ) Protein Plays a Major Role in Aβ-Induced Alterations of Glutamatergic Synaptic Transmission and Plasticity. The Journal of Neuroscience 34, 12893, doi:10.1523/JNEUROSCI.1201-14.2014 (2014).

17 Haque, M. A., Hossain, M. S., Bilkis, T., Islam, M. I. & Park, I.-S. Evidence for a Strong Relationship between the Cytotoxicity and Intracellular Location of β-Amyloid. Life 12 (2022).

18 Kobro-Flatmoen, A., Nagelhus, A. & Witter, M. P. Reelin-immunoreactive neurons in entorhinal cortex layer II selectively express intracellular amyloid in early Alzheimer’s disease. Neurobiology of Disease 93, 172–183, 10.1016/j.nbd.2016.05.012 (2016).

19 Lee, J.-H. et al. Faulty autolysosome acidification in Alzheimer’s disease mouse models induces autophagic build-up of Aβ in neurons, yielding senile plaques. Nature Neuroscience 25, 688–701, doi:10.1038/s41593-022-01084-8 (2022).

20 Suelves, N. et al. Senescence-related impairment of autophagy induces toxic intraneuronal amyloid-β accumulation in a mouse model of amyloid pathology. Acta Neuropathol Commun 11, 82, doi:10.1186/s40478-023-01578-x (2023).

21 Zhang, W. et al. Impairment of the autophagy–lysosomal pathway in Alzheimer’s diseases: Pathogenic mechanisms and therapeutic potential. Acta Pharmaceutica Sinica B 12, 1019–1040, 10.1016/j.apsb.2022.01.008 (2022).

22 Sanchez-Varo, R. et al. Abnormal accumulation of autophagic vesicles correlates with axonal and synaptic pathology in young Alzheimer’s mice hippocampus. Acta Neuropathologica 123, 53–70, doi:10.1007/s00401-011-0896-x (2012).

23 Pickford, F. et al. The autophagy-related protein beclin 1 shows reduced expression in early Alzheimer disease and regulates amyloid β accumulation in mice. The Journal of Clinical Investigation 118, 2190–2199, doi:10.1172/JCI33585 (2008).

24 Gyure, K. A., Durham, R., Stewart, W. F., Smialek, J. E. & Troncoso, J. C. Intraneuronal Aβ-Amyloid Precedes Development of Amyloid Plaques in Down Syndrome. Archives of Pathology & Laboratory Medicine 125, 489–492, doi:10.5858/2001-125-0489-IAAPDO (2001).

25 Mori, C. et al. Intraneuronal Abeta42 accumulation in Down syndrome brain. Amyloid 9(**2**), 88–102 (2002).

26 Oddo, S., Caccamo, A., Smith, I. F., Green, K. N. & LaFerla, F. M. A Dynamic Relationship between Intracellular and Extracellular Pools of A&#x3b2. The American Journal of Pathology 168, 184–194, doi:10.2353/ajpath.2006.050593 (2006).

27 Ismail, R. et al. The relationships between neuroinflammation, beta-amyloid and tau deposition in Alzheimer’s disease: a longitudinal PET study. Journal of Neuroinflammation 17, 151, doi:10.1186/s12974-020-01820-6 (2020).

28 Wright, A. L. et al. Neuroinflammation and Neuronal Loss Precede Aβ Plaque Deposition in the hAPP-J20 Mouse Model of Alzheimer’s Disease. PLOS ONE 8, e59586, doi:10.1371/journal.pone.0059586 (2013).

29 Hong, S., Dissing-Olesen, L. & Stevens, B. New insights on the role of microglia in synaptic pruning in health and disease. Current Opinion in Neurobiology 36, 128–134, 10.1016/j.conb.2015.12.004 (2016).

30 Hong, S. et al. Complement and microglia mediate early synapse loss in Alzheimer mouse models. Science 352, 712–716, doi:10.1126/science.aad8373 (2016).

31 Jansen, I. E. et al. Genome-wide meta-analysis identifies new loci and functional pathways influencing Alzheimer’s disease risk. Nature Genetics 51, 404–413, doi:10.1038/s41588-018-0311-9 (2019).

32 Guerreiro, R. et al. TREM2 Variants in Alzheimer’s Disease. New England Journal of Medicine 368, 117–127, doi:10.1056/NEJMoa1211851 (2012).

33 Welikovitch, L. A. et al. Early intraneuronal amyloid triggers neuron-derived inflammatory signaling in APP transgenic rats and human brain. Proceedings of the National Academy of Sciences 117, 6844–6854, doi:10.1073/pnas.1914593117 (2020).

34 Liddelow, S. A. et al. Neurotoxic reactive astrocytes are induced by activated microglia. Nature 541, 481–487, doi:10.1038/nature21029 (2017).

35 Fancy, N. et al. Mechanisms contributing to differential genetic risks for TREM2 R47H and R62H variants in Alzheimer’s Disease. medRxiv, 2022.2007.2012.22277509, doi:10.1101/2022.07.12.22277509 (2022).

36 Franjic, D. et al. Molecular Diversity Among Adult Human Hippocampal and Entorhinal Cells. bioRxiv, 2019.2012.2031.889139, doi:10.1101/2019.12.31.889139 (2020).

37 Hodge, R. D. et al. Conserved cell types with divergent features in human versus mouse cortex. Nature 573, 61–68, doi:10.1038/s41586-019-1506-7 (2019).

38 Mathys, H. et al. Single-cell transcriptomic analysis of Alzheimer’s disease. Nature 570, 332–337, doi:10.1038/s41586-019-1195-2 (2019).

39 Lake, B. B. et al. Integrative single-cell analysis of transcriptional and epigenetic states in the human adult brain. Nat Biotechnol 36, 70–80, doi:10.1038/nbt.4038 (2018).

40 Lake, B. B. et al. Neuronal subtypes and diversity revealed by single-nucleus RNA sequencing of the human brain. Science 352, 1586, doi:10.1126/science.aaf1204 (2016).

41 Otero-Garcia, M. et al. Molecular signatures underlying neurofibrillary tangle susceptibility in Alzheimer’s disease. Neuron, 10.1016/j.neuron.2022.06.021 (2022).

42 Bortolomeazzi, M. et al. A SIMPLI (Single-cell Identification from MultiPLexed Images) approach for spatially-resolved tissue phenotyping at single-cell resolution. Nature Communications 13, 781, doi:10.1038/s41467-022-28470-x (2022).

43 Palomero-Gallagher, N. & Zilles, K. Cortical layers: Cyto-, myelo-, receptor- and synaptic architecture in human cortical areas. NeuroImage 197, 716–741, 10.1016/j.neuroimage.2017.08.035 (2019).

44 Schneider, V. A. et al. Evaluation of GRCh38 and de novo haploid genome assemblies demonstrates the enduring quality of the reference assembly.

45 Wonders, C. P. & Anderson, S. A. The origin and specification of cortical interneurons. Nat Rev Neurosci 7, 687–696, doi:10.1038/nrn1954 (2006).

46 Verkhratsky, A., Zorec, R. & Parpura, V. Stratification of astrocytes in healthy and diseased brain. Brain Pathology 27, 629–644, 10.1111/bpa.12537 (2017).

47 Otero-Garcia, M. et al. Molecular signatures underlying neurofibrillary tangle susceptibility in Alzheimer’s disease. Neuron 110, 2929–2948.e2928, 10.1016/j.neuron.2022.06.021 (2022).

48 Langseth, C. M. et al. Comprehensive in situ mapping of human cortical transcriptomic cell types. Communications Biology 4, 998, doi:10.1038/s42003-021-02517-z (2021).

49 Michaki, V. et al. Down-regulation of the ATP-binding Cassette Transporter 2 (Abca2) Reduces Amyloid-β Production by Altering Nicastrin Maturation and Intracellular Localization*. Journal of Biological Chemistry 287, 1100–1111, 10.1074/jbc.M111.288258 (2012).

50 Lee, D.-S., Tomita, S., Kirino, Y. & Suzuki, T. Regulation of X11L-dependent Amyloid Precursor Protein Metabolism by XB51, a Novel X11L-binding Protein*. Journal of Biological Chemistry 275, 23134–23138, 10.1074/jbc.C000302200 (2000).

51 Jungsu, K. et al. BRI2 (*ITM2b*) Inhibits Aβ Deposition *In Vivo*. The Journal of Neuroscience 28, 6030, doi:10.1523/JNEUROSCI.0891-08.2008 (2008).

52 Schnöder, L. et al. Deficiency of Neuronal p38α MAPK Attenuates Amyloid Pathology in Alzheimer Disease Mouse and Cell Models through Facilitating Lysosomal Degradation of BACE1*. Journal of Biological Chemistry 291, 2067–2079, 10.1074/jbc.M115.695916 (2016).

53 Burbulla, L. F. et al. Dissecting the role of the mitochondrial chaperone mortalin in Parkinson’s disease: functional impact of disease-related variants on mitochondrial homeostasis. Hum Mol Genet 19, 4437–4452, doi:10.1093/hmg/ddq370 (2010).

54 Jordi, M., Roy, C. S., Kenneth, W. & Henry, W. Q. Heat Shock Protein 70 Participates in the Neuroprotective Response to Intracellularly Expressed β-Amyloid in Neurons. The Journal of Neuroscience 24, 1700, doi:10.1523/JNEUROSCI.4330-03.2004 (2004).

55 Bordi, M. et al. A gene toolbox for monitoring autophagy transcription. Cell Death & Disease 12, 1044, doi:10.1038/s41419-021-04121-9 (2021).

56 Fleming, A. et al. The different autophagy degradation pathways and neurodegeneration. Neuron 110, 935–966, 10.1016/j.neuron.2022.01.017 (2022).

57 Jessica, E. R. et al. Disease-specific selective vulnerability and neuroimmune pathways in dementia revealed by single cell genomics. bioRxiv, 2023.2009.2029.560245, doi:10.1101/2023.09.29.560245 (2023).

58 Crooke, P. S., III, Tossberg, J. T., Heinrich, R. M., Porter, K. P. & Aune, T. M. Reduced RNA adenosine-to-inosine editing in hippocampus vasculature associated with Alzheimer’s disease. Brain Communications 4, fcac238, doi:10.1093/braincomms/fcac238 (2022).

59 Gómez-Isla, T. et al. Neuronal loss correlates with but exceeds neurofibrillary tangles in Alzheimer’s disease. Annals of Neurology 41, 17–24, 10.1002/ana.410410106 (1997).

60 Moloney, C. M., Lowe, V. J. & Murray, M. E. Visualization of neurofibrillary tangle maturity in Alzheimer’s disease: A clinicopathologic perspective for biomarker research. Alzheimer’s & Dementia 17, 1554–1574, 10.1002/alz.12321 (2021).

61 Gratuze, M., Leyns, C. E. G. & Holtzman, D. M. New insights into the role of TREM2 in Alzheimer’s disease. Molecular Neurodegeneration 13, 66, doi:10.1186/s13024-018-0298-9 (2018).

62 Konishi, H. et al. Astrocytic phagocytosis is a compensatory mechanism for microglial dysfunction. The EMBO Journal 39, e104464, 10.15252/embj.2020104464 (2020).

63 Damisah, E. C. et al. Astrocytes and microglia play orchestrated roles and respect phagocytic territories during neuronal corpse removal in vivo. Science Advances 6, eaba3239, doi:10.1126/sciadv.aba3239.

64 Wood, J. I. et al. Plaque contact and unimpaired Trem2 is required for the microglial response to amyloid pathology. Cell Reports 41, 111686, 10.1016/j.celrep.2022.111686 (2022).

65 D’Andrea, M. R., Nagele, R. G., Wang, H. Y., Peterson, P. A. & Lee, D. H. S. Evidence that neurones accumulating amyloid can undergo lysis to form amyloid plaques in Alzheimer’s disease. Histopathology 38, 120–134, 10.1046/j.1365-2559.2001.01082.x (2001).

66 Piras, A., Collin, L., Grüninger, F., Graff, C. & Rönnbäck, A. Autophagic and lysosomal defects in human tauopathies: analysis of post-mortem brain from patients with familial Alzheimer disease, corticobasal degeneration and progressive supranuclear palsy. Acta Neuropathol Commun 4, 22, doi:10.1186/s40478-016-0292-9 (2016).

67 Perez-Nievas, B. G. & Serrano-Pozo, A. Deciphering the Astrocyte Reaction in Alzheimer’s Disease. Frontiers in Aging Neuroscience 10 (2018).

68 Wei, S. et al. SOX9 Is an Astrocyte-Specific Nuclear Marker in the Adult Brain Outside the Neurogenic Regions. The Journal of Neuroscience 37, 4493, doi:10.1523/JNEUROSCI.3199-16.2017 (2017).

69 Kecheliev, V. et al. Aquaporin 4 is differentially increased and dislocated in association with tau and amyloid-beta. Life Sciences 321, 121593, 10.1016/j.lfs.2023.121593 (2023).

70 Dai, D. L., Li, M. & Lee, E. B. Human Alzheimer’s disease reactive astrocytes exhibit a loss of homeostastic gene expression. Acta Neuropathol Commun 11, 127, doi:10.1186/s40478-023-01624-8 (2023).

71 Youmans, K. L. et al. Intraneuronal Aβ detection in 5xFAD mice by a new Aβ-specific antibody. Molecular Neurodegeneration 7, 8, doi:10.1186/1750-1326-7-8 (2012).

72 Shabani, K. et al. The temporal balance between self-renewal and differentiation of human neural stem cells requires the amyloid precursor protein. Science Advances 9, eadd5002, doi:10.1126/sciadv.add5002.

